# Comprehensive Understanding of Hydrogeochemical evaluation of seasonal variability in groundwater quality Dynamics in the Gold Mining Areas of Osun State, Nigeria

**DOI:** 10.1101/2022.11.06.22282015

**Authors:** Awogbami Stephen Olalekan, Solomon Olayinka Adewoye, Sawyerr Olawale Henry, Opasola Afolabi Olaniyi, Morufu Olalekan Raimi

## Abstract

**Background:** A crucial issue directly affecting the wellbeing of the human race is water quality. Within a few decades, a greater reliance on groundwater was needed to supply water for drinking, agriculture, and other uses due to the scarcity and contamination of surface water. To forecast its long-term use and increase output, irrigated agriculture requires high-quality water, which must be measured.

**Objective:** The goals of the current study are to comprehend the hydro-geochemistry, drinking water appropriateness, and occurrence of hydro-geochemistry concepts in the groundwater of the gold mining regions of Osun State, South-West Nigeria.

**Method:** Standardized analytical methods were used in the research. All sampling, conservation, transportation, and analysis were carried out in accordance with APHA guidelines (2012). To prevent deterioration of the organic compounds, all acquired samples were sent to research laboratory, while maintaining in an icebox.

**Results:** The study also identifies critical pollutants, affecting the ground water quality within its course through the gold mining areas of Osun State. Finally, Colour, pH, DO, EC, TDS, TSS, TS, Hardness, Magnesium, Nitrate, Phosphate, Lead, Cadmium, Chromium, Manganese, Mercury and Arsenic have been found to be critical parameters for the stretch in each season of this research.

**Conclusion:** The authors advise developing ongoing systems for monitoring water quality as well as efficient management techniques to prevent excessive groundwater pollution. These findings will therefore aid decision-makers in managing pollution in Osun State’s gold mining zones and better understanding the impact of different seasons on water quality. The findings of this study can serve as a foundation for the future monitoring of the effects of anthropogenic activities on local watercourses when mining companies are developed. This requires consideration in terms of both regulatory measures and proactive initiatives for addressing the ensuing issues in the future. In order to maintain sustainability, a long-term monitoring plan is suggested in this study to be implemented into the groundwater bodies to restore their quality.

## 1. Introduction

These days, among the most significant environmental problems worldwide are the buildup of heavy metals and the poisoning of groundwater, particularly agricultural soils, both of which are mostly brought on by human activity. The mining sector is a vital pillar of social development and a significant portion of the national economy [1-23]. Despite producing billions of tons of tailings waste each year, mining has enormous social and economic advantages [24]. Massive mine waste is typically heaped in an untreated open tailings pond. They take up a lot of space and are full of dangerous heavy metals that could be harmful to both individuals and the environment [23]. According to earlier research, tailings ponds contaminate the soil, dust, water, sediments, and crops in the vicinity with heavy metals. Heavy-metal contamination of agricultural soils has become a significant environmental concern in human cultures, particularly in developing nations, at the same time as the growth and expansion of industry, mining operations, and excessive use of chemical fertilizers [25-35]. Studies have demonstrated that anthropogenic activities such as improper waste management [36-37], industrial, mining, agricultural, and domestic activities as well as geogenic processes like weathering of rock minerals, rainfall intensity, leaching, and local geology of the aquifer [1-13] are the major controls on groundwater quality. Groundwater was severely contaminated as a result of excessive and unsustainable groundwater extraction for drinking and irrigation, irrigation runoff, surface contaminants permeating the groundwater system, a high rate of evaporation, etc. [7–13]. In addition to these surface contaminants, groundwater quality was further impacted by geochemical processes, land use, and regional geology [38, 39].

Therefore, thorough understanding of the hydrogeochemical properties, geogenic, and anthropogenic processes regulating groundwater chemistry are essential to solving groundwater-related problems. By establishing the drinking and irrigation appropriateness of groundwater to maintain human health while increasing agricultural production, numerous researchers from around the world have made contributions to this topic [1-35]. Numerous studies have been conducted to understand how the groundwater system behaves in the context of oil and gas mining, both under natural and anthropogenic drives [1–12]. For example, Raimi and Sawyerr [11] studied the preliminary study of groundwater quality using hierarchical classification approaches for contaminated sites in indigenous communities associated with crude oil exploration facilities in Rivers State and concluded that the findings of this study are significant for policymakers and agencies in terms of dealing with the issues identified to enhance sustainable livelihood practices in the oil rich Niger Delta region of Nigeria. Therefore, decision-makers should take proper initiatives to get local people aware of the endangered zones before use, as drinking water is key to good health. Similarly, multinational oil companies will find it useful in their quest for viable social corporate responsibility and remediation plans in their respective host communities. The method proved to be a useful and objective tool for environmental planning. Raimi *et al.* [9] expound the hydrogeochemical and multivariate statistical techniques to trace the sources of ground water contaminants and affecting factors of groundwater pollution in an oil and gas producing wetland in Rivers State, the study conclude that greater efforts should be made to safeguard groundwater, which is hampered by geogenic and anthropogenic activities, in order to achieve sustainable groundwater development. As a result, communities are recommended to maintain a groundwater management policy to ensure long-term sustainability. The study is useful for understanding groundwater trace sources in Rivers State’s Ebocha-Obrikom districts. Such understanding would enable informed mitigation or eradication of the possible detrimental health consequences of this groundwater, whether through its use as drinking water or indirectly through consumption of groundwater-irrigated crops. As a result, determining its primary probable source of pollution (MPSP) is critical since it provides a clearer and more immediate interpretation. Furthermore, the research findings can be used as a reference for groundwater pollution prevention and water resource protection in the Niger Delta region of Nigeria. Also, Olalekan *et al.,* [10] (2022) studied quality water, not everywhere: assessing the hydrogeochemistry of water quality across Ebocha-Obrikom Oil and Gas Flaring Area in the Core Niger Delta Region of Nigeria and concluded that it is recommended that the local government environmental health officers and other regulatory agencies frequently monitor the levels of these pollutants within the area and ensure strict adherence to guidelines to ensure a healthy environment. As exposure to the parameters can have a remarkable impact on human health living in the vicinity of the gas flaring area through drinking water around the study area, groundwater needs to be treated prior to any use for household or drinking purposes. Thus, this study would help in decision making of the stakeholders and relevant authorities for execution of reasonable groundwater management strategies and remediation plans in the area to protect public and environmental health. Raimi *et al.,* [2] toxicants in water: hydrochemical appraisal of toxic metals concentration and seasonal v variation in drinking water quality in oil and gas field area of Rivers State, the study concluded that extra efforts must be made to completely understand the hydrogeochemical properties and appropriateness of groundwater in Nigeria’s core Niger Delta region in order to ensure quality groundwater supply for varied applications. As a result, this study will help build a quantitative understanding of the effects of many causes on variations in groundwater levels in every aquifer on the planet. This analysis also supports a valuable tool for academics, activists, and public servants who want to raise community awareness and enhance performance. The judgments will be an invaluable resource for policymakers, the Ministry of Water Resources, and development professionals because they emphasize the need for appropriate methods to reduce toxic contamination of water resources in the central Niger Delta in order to safeguard the public health from risks that could be both carcinogenic and non-carcinogenic. Because of the growth in population, urbanization, and industrialization, the issue of environmental pollution is getting worse and worse every day around the world [40–51]. Hazardous pollution contamination of soil [17], sediments [23, 24], surface water [6, 13], groundwater [1–5], and air [52–55] is an issue on a global scale. The most dangerous group of pollutants are heavy metals, which include cadmium (Cd), nickel (Ni), lead (Pb), aluminum (Al), arsenic (As), mercury (Hg), lithium (Li), copper (Cu), silver (Ag), and zinc (Zn). This is because these metals accumulate easily in the food chain and are persistent by nature [1–5]. In mining and agricultural towns in Nigeria, there have been concerns about the quality of the groundwater. The majority of this groundwater, according to reports, is unfit for drinking and irrigation, however some may be suitable for industrial uses. According to Raimi et al. [12], background values for various elements are assessed either spatially or temporally (concentrations prior to anthropogenic activity) (concentrations in the areas not influenced by anthropogenic activity). Since groundwater is essential for about 75% of Nigeria’s irrigated agriculture, groundwater is the main factor causing this development [56], apart from this, 85% of drinking water supplies are also met by groundwater. This over reliance on groundwater causes soil deterioration, water pollution, seawater intrusion, and groundwater level decline [17, 47]. Global, regional, and local levels of understanding on groundwater are all lacking [18]. As a result, there is an increasing need to address the concerns with groundwater quality because poor water quality has an impact on both crop output and human health. Anthropogenic and geological factors have a significant impact on changes in groundwater quality. If anthropogenic activities are to blame for the deterioration, environmental measures can preserve the groundwater [57–63]. However, it poses a significant problem if geogenic sources have an impact on groundwater quality. Anthropogenic factors like salinity (because to over-extraction in coastal aquifers) and nitrates (due to agricultural runoff) as well as geogenic factors like fluoride, arsenic, and iron significantly affect groundwater in many areas of the Niger Delta. The focus of the current study is on the seasonal variation of ground water’s hydro-geochemistry since iron and fluoride, out of all the contaminants, pose the greatest hazard to public health in Nigeria [1–12]. The current study’s goals are to comprehend the hydro-geochemistry of the groundwater in the mining region; evaluate the groundwater’s appropriateness for drinking and irrigation; and comprehend the presence of physico-chemical and heavy metals in groundwater. In order to measure the impact of anthropogenic and geogenic processes on groundwater quality while projecting future availability, the integrated hydrochemical assessment of groundwater is necessary. In order to manage soil resources sustainably and avoid any risks to human health, it is also helpful, as this could ultimately increase agricultural production [17]. Because ground water resources are so crucial to both human health and natural ecosystems, it is extremely important to monitor both their qualitative and quantitative changes over time [2–5].

The monitoring of groundwater quality during the last few decades has increased by means of measurement of various water quality indicators. Nonetheless, a monitoring program that offers a representative and accurate prediction of ground waters quality is required due to geographical and temporal fluctuations in water quality, which are frequently challenging to interpret. The majority of Osun State is made up of agricultural land, with groundwater serving as the main source of drinking water and irrigation for the state’s gold mining regions. Agriculture is a major industry in this region. The results of this study will undoubtedly benefit sustainable groundwater resource management, groundwater quality improvement, and the avoidance of potential health risks.

## 2. Material and Methods

### The Study Area

Surface water samples analyzed in this research were collected for twelve (12) months (June 2020 to May 2021) from three surface water bodies from different locations. The surface water bodies include Aye-Oba River, Alapadi and Eti-oni stream; both Alapadi and Eti-oni stream are tributaries of Aye-oba River. All situated within gold mining activities areas across three Local Governments (Ife South, Atakumosa East, and Atakumosa West) in Osun State, Southwest Nigeria. Aye-Oba River lies between the latitude 7° 23’ N and longitude of 4° 61^’^ E (see figure 1-5). The river is very significant to the socio-economic growth of the residents of Ife South Local Government area in Osun State to sustain subsistence farming such as irrigation, livestock, and fishery. The river is the major source of water supply for consumption and domestic use. The river was constructed as a dam in the year 2004. Thus, the dam served as the major source of potable water supply to more than fifteen communities in Ife South Local Government areas and its environs until it stopped the operation (figure 1-5).

**Fig. 1:**
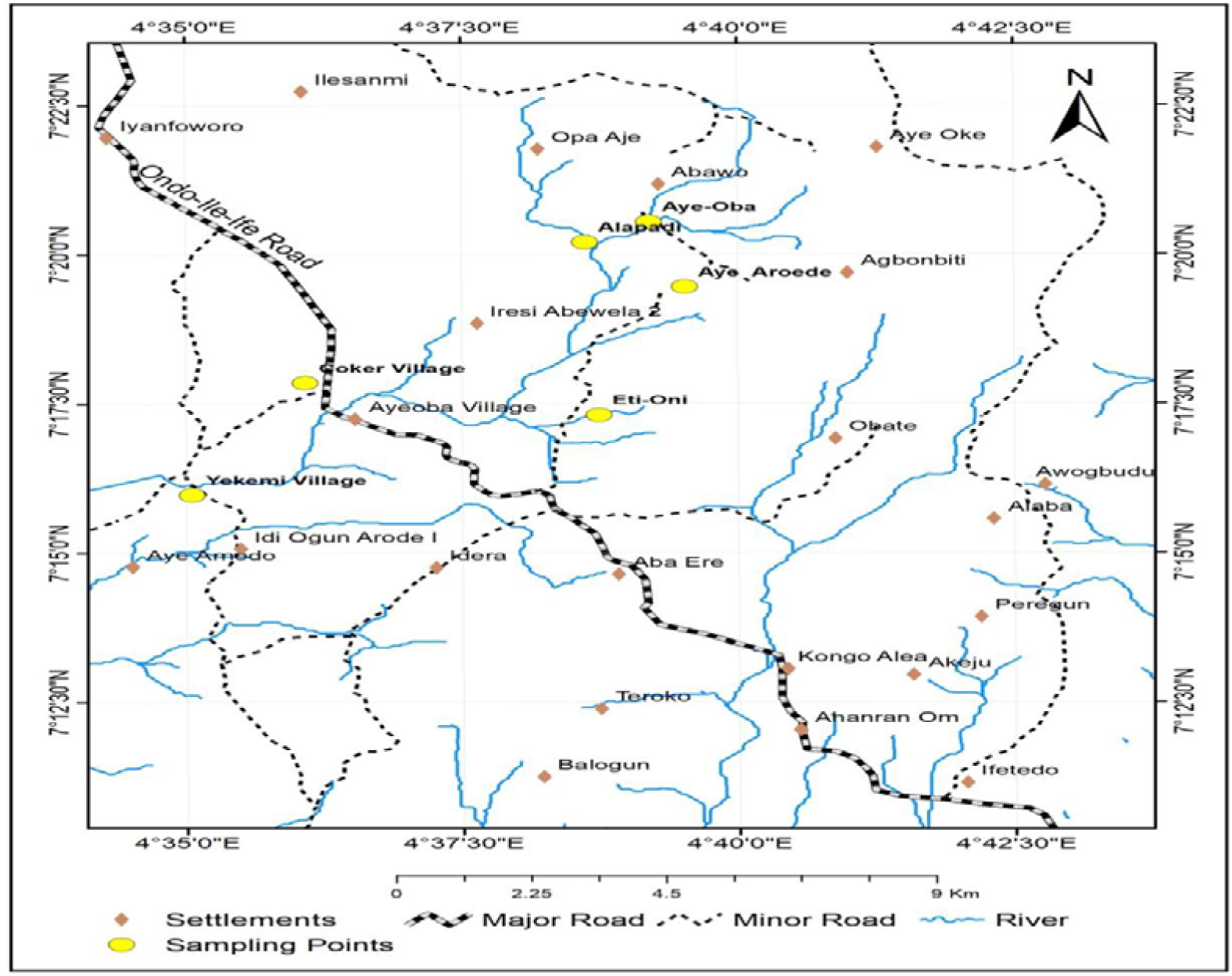
Area map showing sampling Aye-Oba River and its tributaries.

**Fig. 2:**
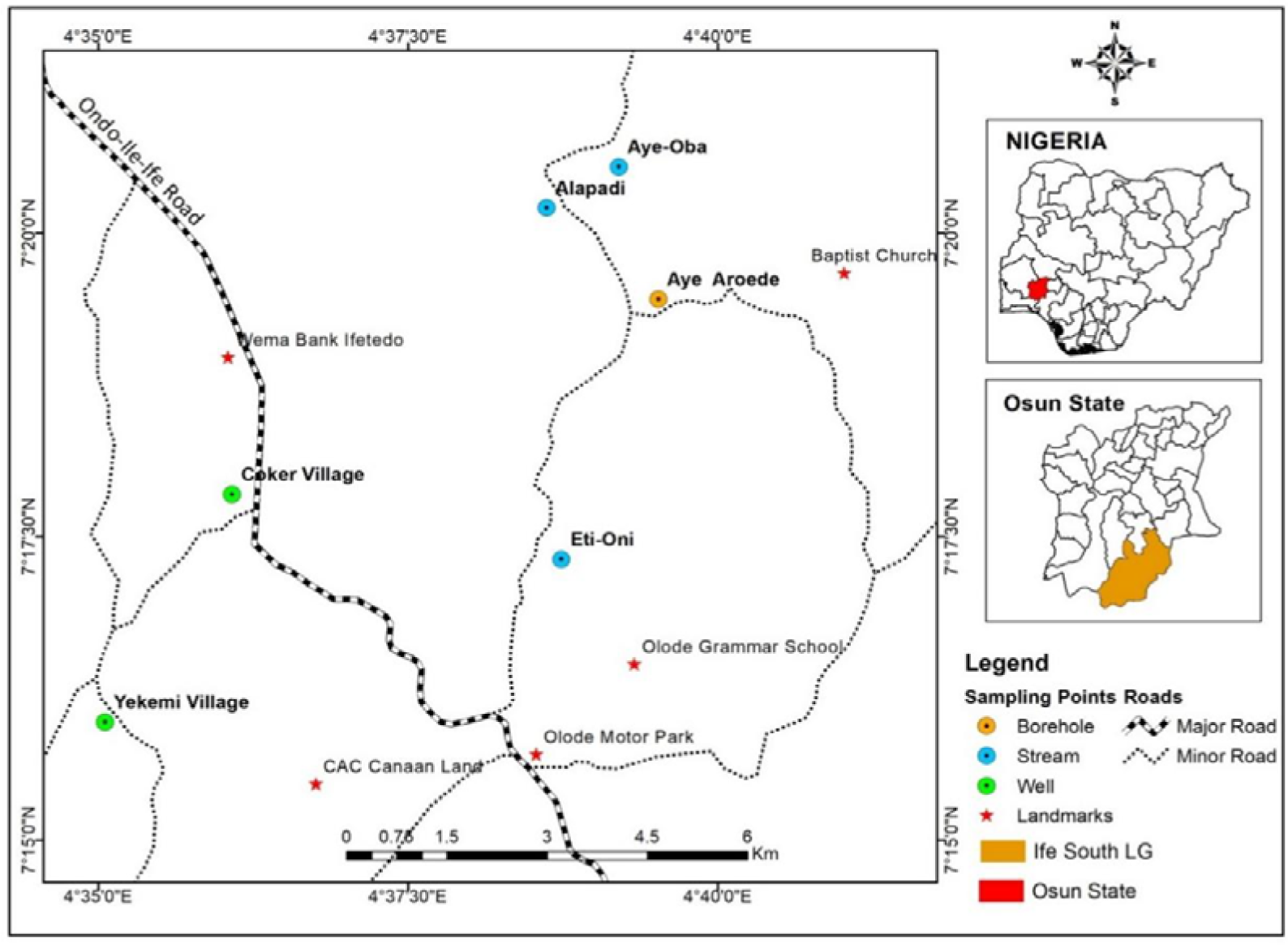
Map showing sampling points at Aye-oba River and in Ife South.

**Fig. 3:**
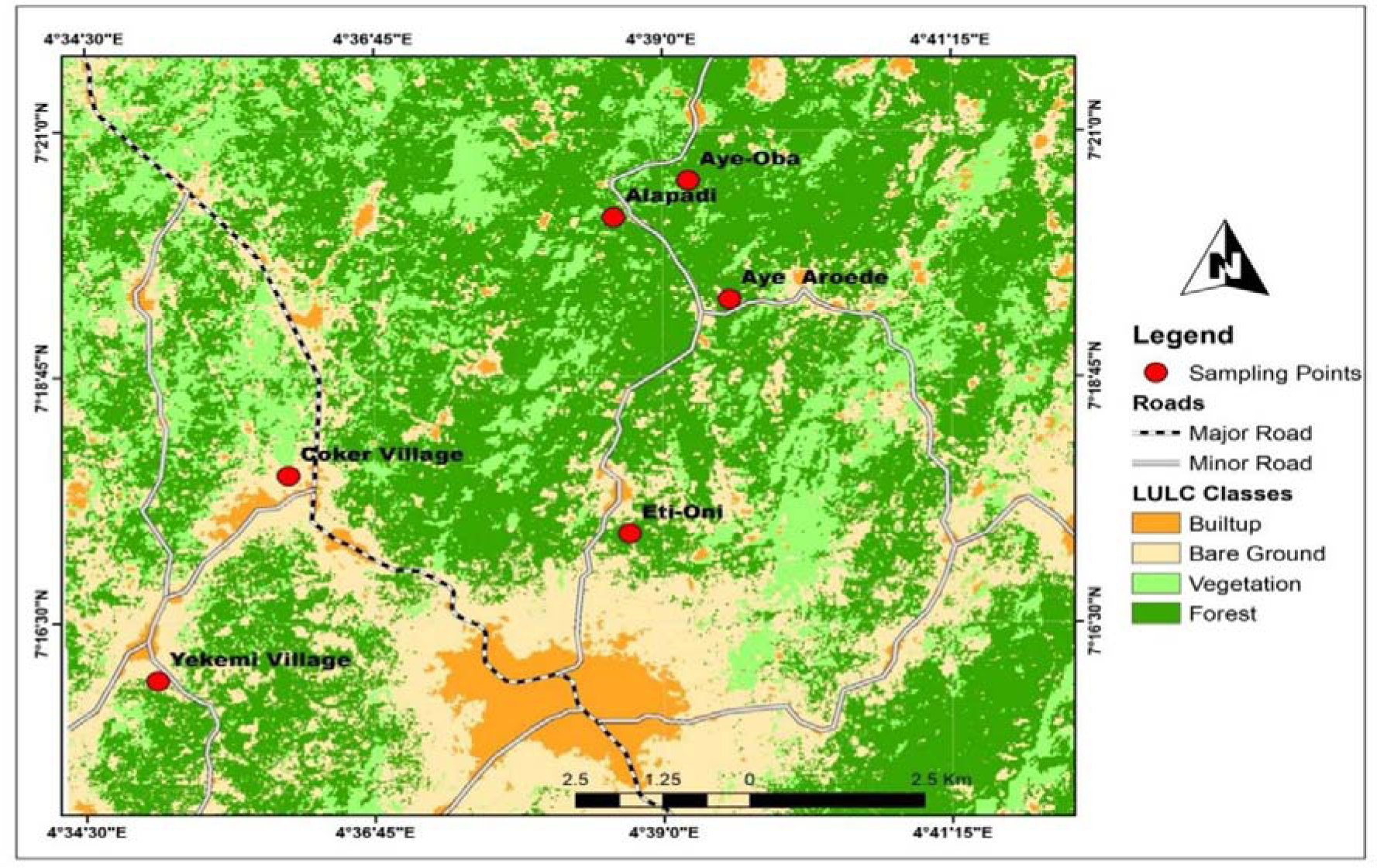
Map showing sampling points and LULC Classes in Ife South.

**Fig. 4:**
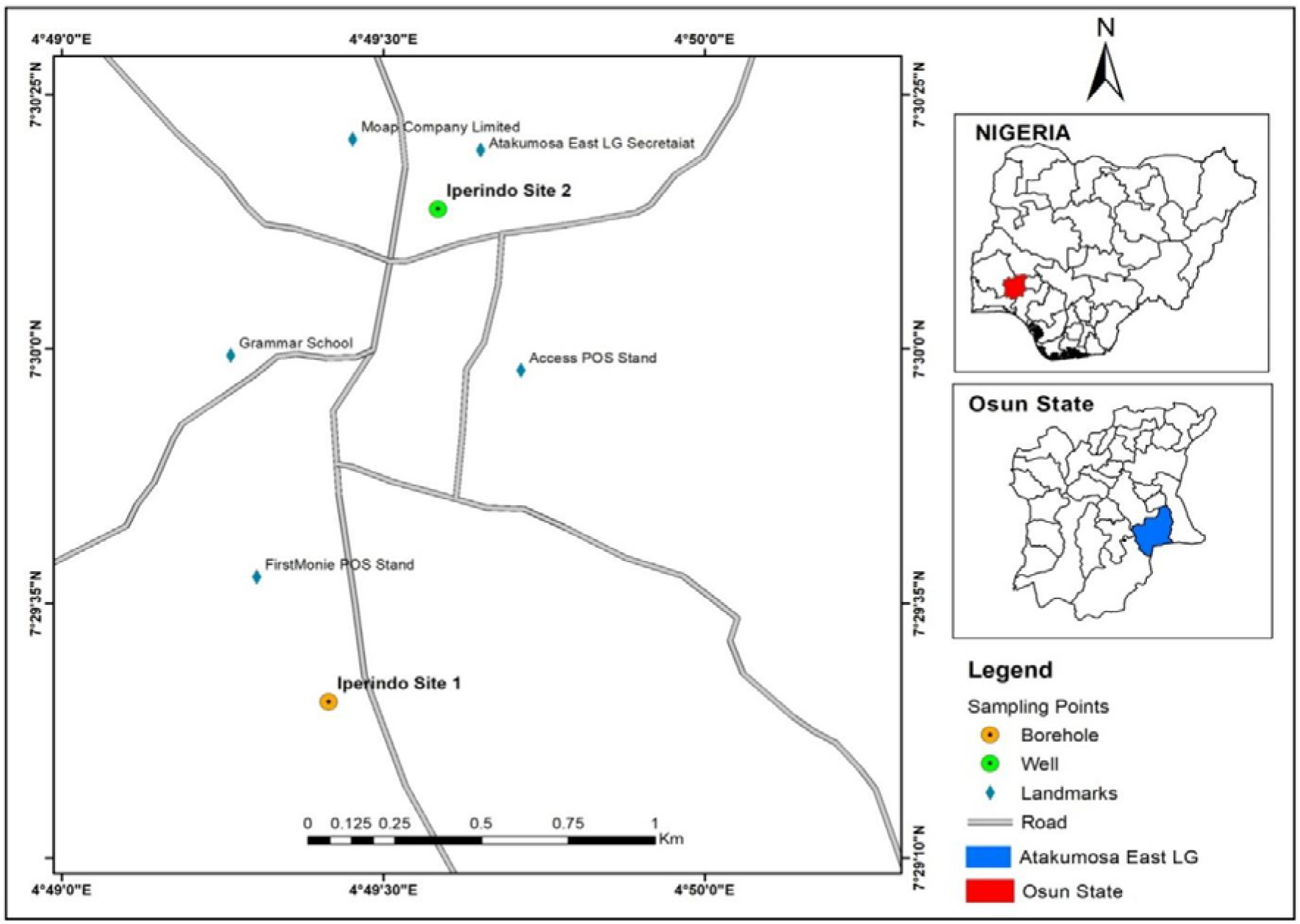
Map showing sampling sites in Atakumosa East.

**Fig. 5:**
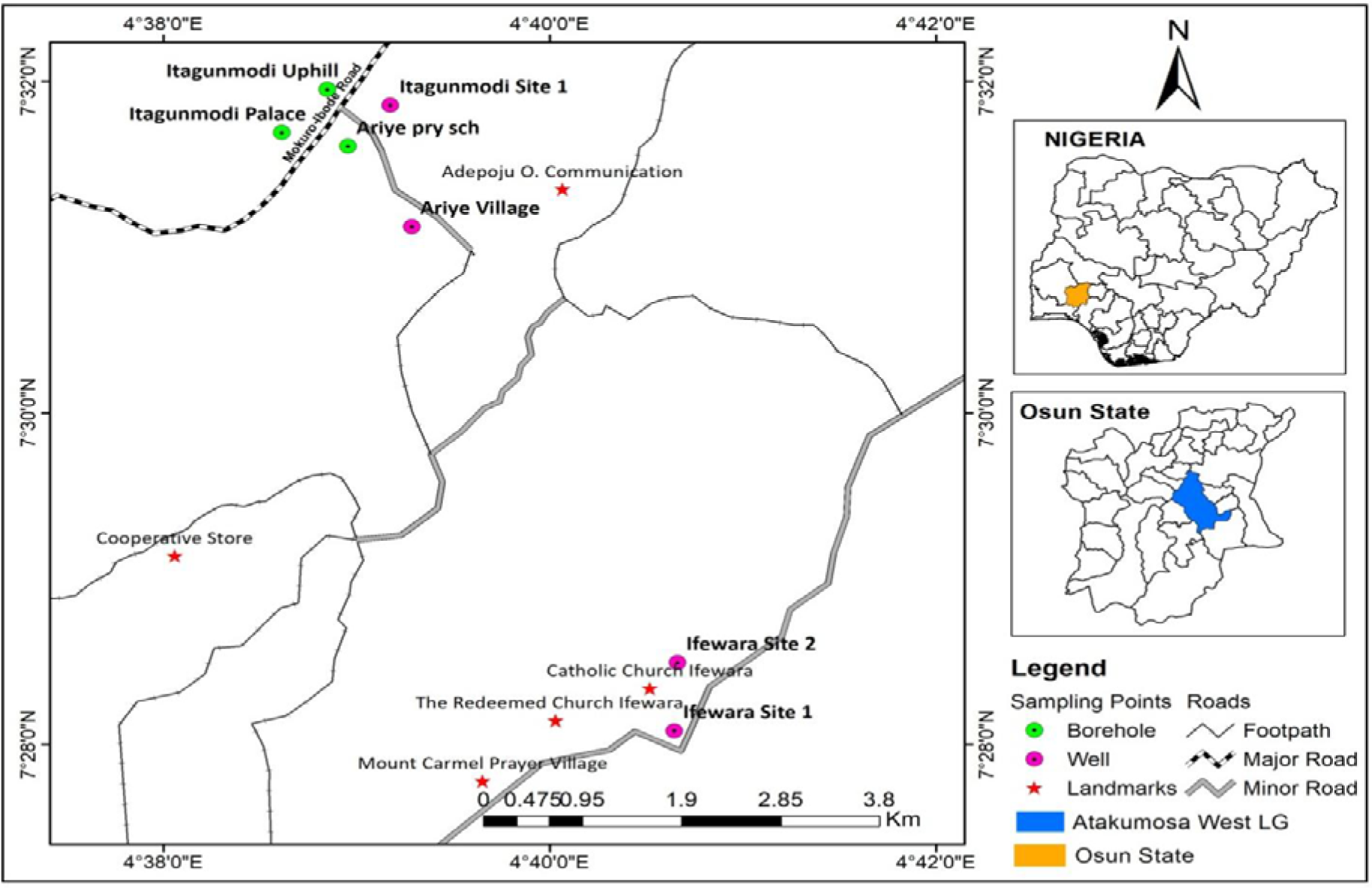
Map showing sampling points in Atakumosa West.

### Experimental design and description of the sampling points

The study design employed was laboratory-based experimental approach and a descriptive cross-sectional study coupled with field observations. The sampling points were: upper stream (site A), middle stream (site B) and lower steam (site C) (see figure 1-5 below). The division was done in relation to the discharge of effluents from mining, domestic and agricultural activities, most especially pesticide pollution through various forms of indiscriminate application on farmlands and domestic effluents particularly from the oil mills processing that enter the stream. Site A (upper stream) represent Eti-oni stream, this stream is about 500 meters from the mining site and about 1km to site B the effluent discharge point. Site B (middle stream) represents Alapadi stream. This stream directly receives effluents discharge due to mining activities in the area. It also receives significant effluents from oil palm processing units located along the river bank and run-off water from farmlands, cassava processes, and refuse dump from homes. Site C (lower stream) represent Aye-Oba River/dam, the abandoned dam. The river is about three (3) kilometers from site B. Physical observation showed that the effluents received by the water bodies from site A and B has changed the colour of the water bodies and there was unpleasant smell emanating from the river. In addition to this, the river receives effluents from cassava processing wastes, open defecation, poultry wastes, animal dung, pesticide products, and water run-off from dumpsite. Therefore, the choice of the afore-mentioned sampling points, presented in figure 1-5, was based on the accessibility, the rate at which they receive effluents from different sources, the extent of their pollution, and particularly their distances from the site of mining activities. In addition, sampling sites were chosen based on the potential exposure of the surface water to different sources of agro-industrial and other sources of pollutants.

### Water sample collection for heavy metal analysis

Collection of water samples was conducted between the hours of 8.00 am and 12 noon, every fourth week of the month in all the sampling points for the period of 12 months (June 2020 to May 2021). Water samples were collected into plastic bottles which were previously soaked in 3% nitric acid and washed with distilled water before sampling, this was in accordance to the method described by APHA [64]. Water samples collected for the determination of dissolved oxygen were collected in dark glass containers and fixed on the spot with Winkler reagent. The water samples were properly preserved following the water sample preservation methods described by APHA [64]. As a result, figure 6 displays the primary approaches for determining the composition of groundwater water.

**Fig. 6:**
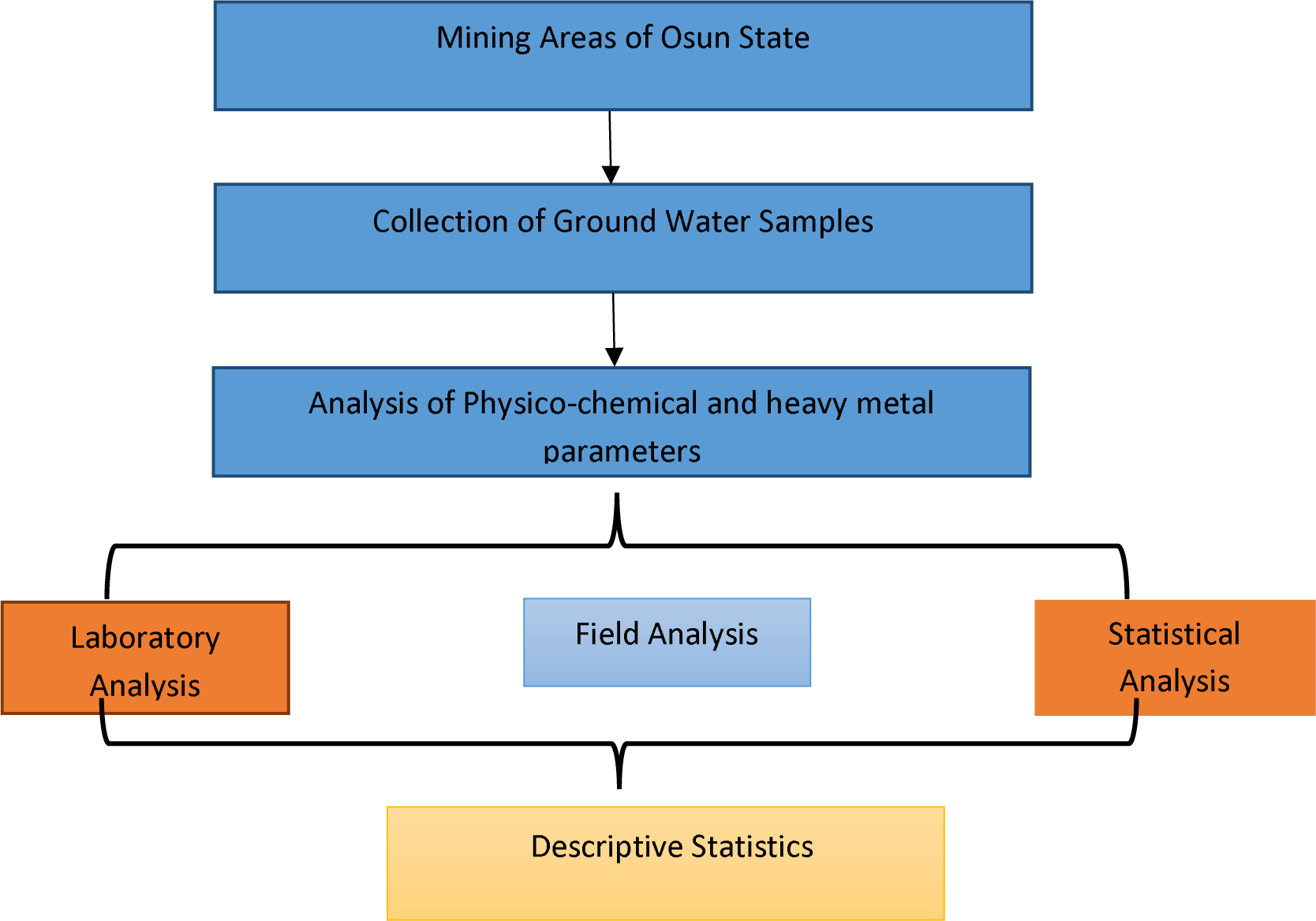
A diagram depicting the quantification methods used in the current investigation. Adapted from Raimi *et al.,* [9], Olalekan *et al.,* [10], Raimi & Sawyerr [11], Raimi *et al.,* [12]

## 3. Results and Discussion

### 3.1. Hydrogeochemistry, suitability and monthly variations in physicochemical composition of groundwater for drinking/irrigation purpose

Figure 7-17 provides a hydrochemical data overview. in the area of study. Because of the growth in population, urbanization, and industrialization, the issue of environmental pollution is getting worse and worse every day around the world [57–63]. Hazardous pollution contamination of groundwater is a concern on a global scale [8–12]. The most dangerous group of pollutants are heavy metals, which include cadmium (Cd), nickel (Ni), lead (Pb), aluminum (Al), arsenic (As), mercury (Hg), lithium (Li), copper (Cu), silver (Ag), and zinc (Zn) [47–49]. These pollutants are easily accumulated in the food chain and are persistent in nature. The outcomes were contrasted with those of the World Health Organization [65] and the Nigerian Standard Organization [66]. The research location was an agricultural province with deficient waste management methods and a gold mining setting. It is thought that a number of contaminants, including agrochemicals and domestic garbage, may have affected the groundwater resource in this area. After that, as locals rely on these water sources for drinking, domestic use, and irrigation, the physicochemical parameters of the groundwater were examined to ascertain the general quality of the water.

**Fig. 7:**
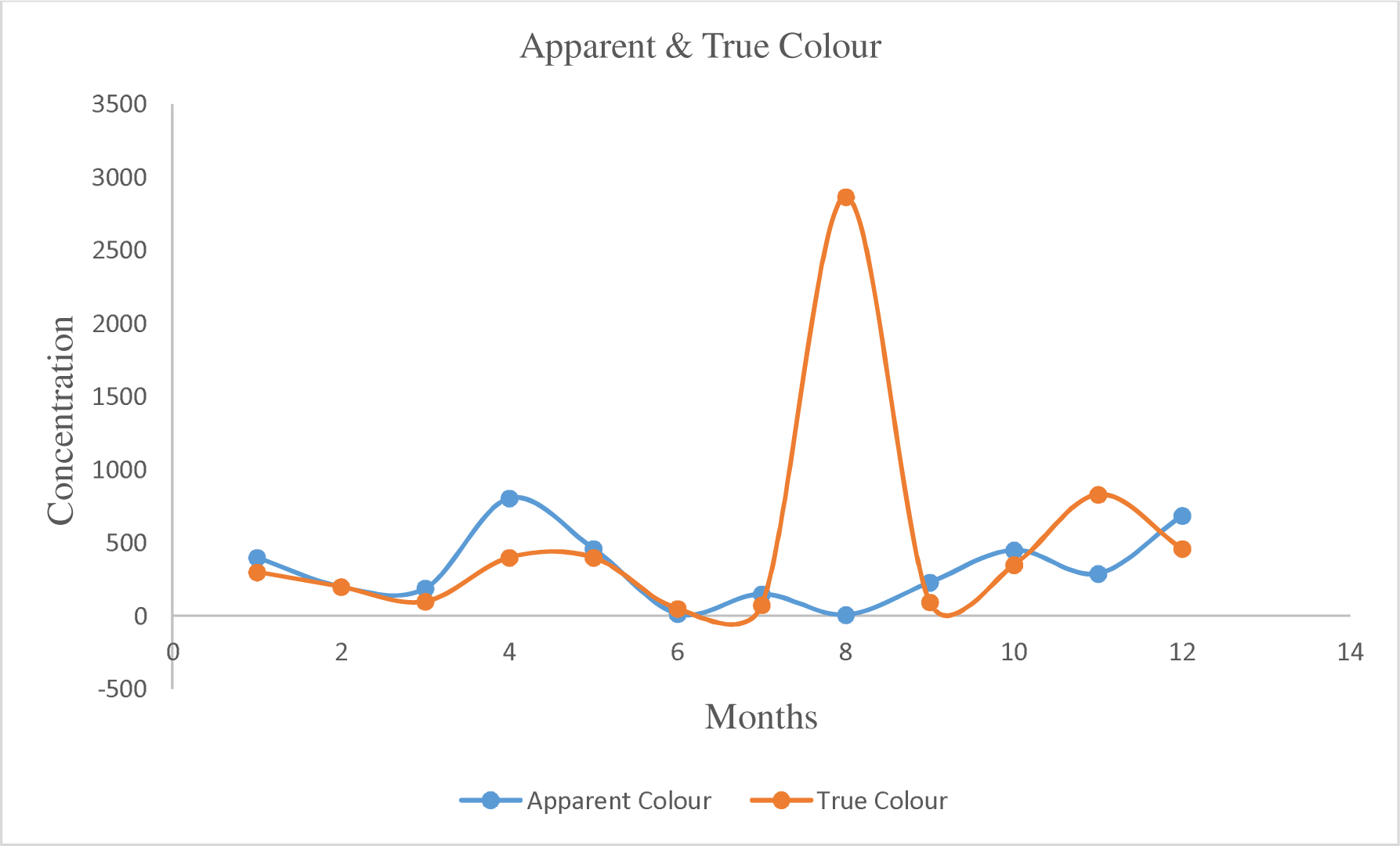
Monthly variation of apparent & true colour in ground water

For apparent colour in general, the monthly variation in apparent colour groundwater is given in Fig. 7. The apparent colour of groundwater was observed to be higher in the month of April (805.01), the onset month of the rainy season and December (686.73) during the dry season. The lowest points were between June and August (8.75 – 150.07) which fall within the period of the rainy season. Hence, any water with a characteristic colour insinuates contamination. While, for true colour, the monthly variations in true colour of groundwater is given in Fig. 7. The values of true colour of groundwater reached its peak in August (2864.9) and reduce drastically in September (95.3); and attained a second smaller peak in November (830.98). The lowest value of groundwater true colour was recorded in the month of June (50.84). Thus, the colour at groundwater in the mining environment could be attributed to dissolved salts and other materials occurring as natural resources as well as contaminants must have influenced colour of the study areas.

The TOC and TOM level in groundwater is given in Fig. 8. The TOC and TOM are scientifically related as the TOM can be estimated from TOC. The peak values of TOC and TOM were recorded between June and July (20.45 - 20.65 mg/L) (35.0mg/L – 35.0mg/L) was recorded respectively. From the month of August (3mg/L), September (5.0)(8.0)mg/L, October (6)(10)mg/L, November (7)(12.5)mg/L, and December (8.5)(15.0)mg/L there is inclination step of the level of TOC and TOM.

**Fig. 8:**
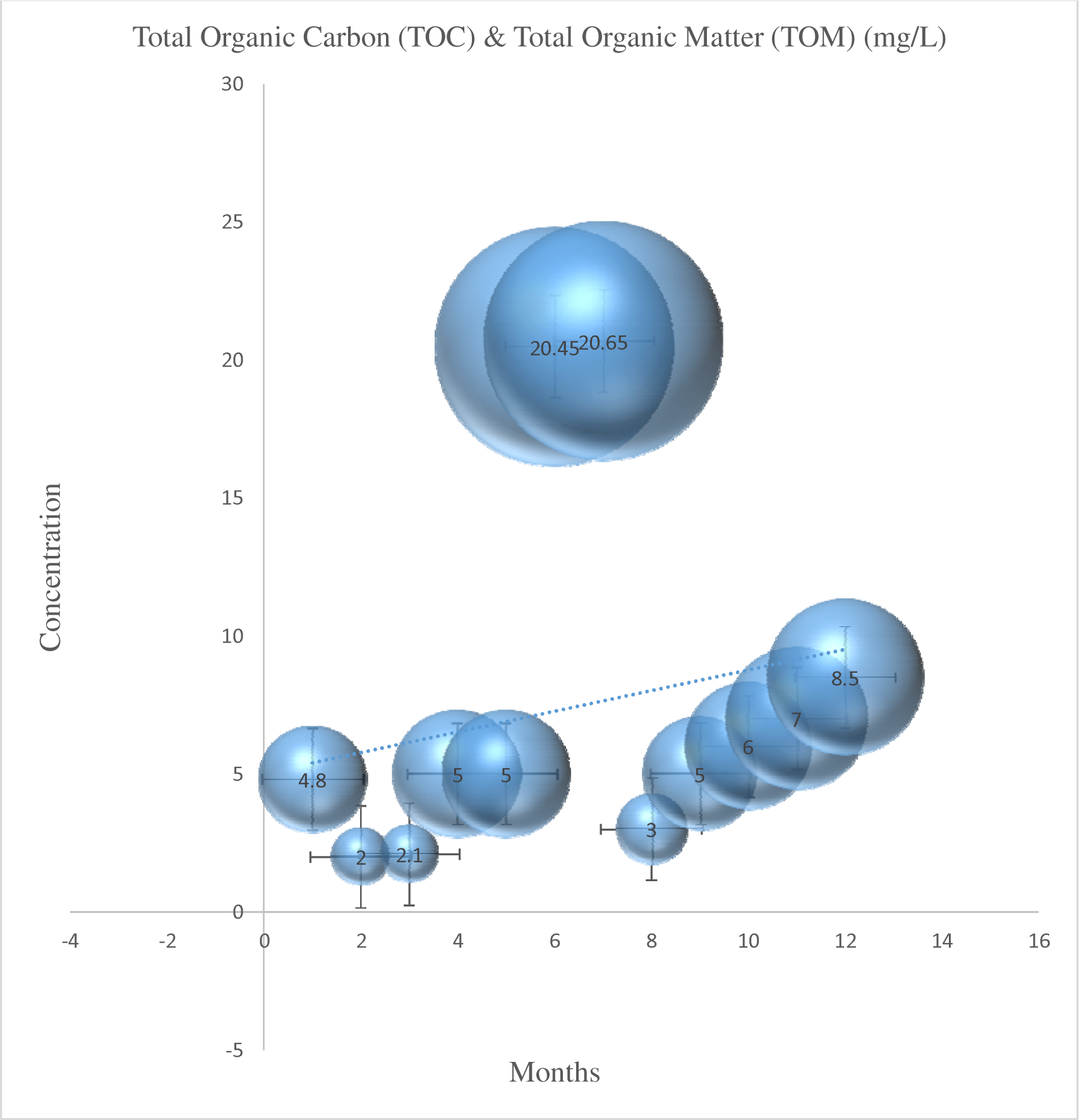
Monthly variation of TOC & TOM in ground water

Biochemical Oxygen Demand is the amount of oxygen needed by bacteria and other microorganisms to break down organic waste in aerobic environments at a particular temperature (BOD). During fermentation, bacteria and other microbes eat organic materials. The broken organics were changed into CO_2_ and H_2_O simple molecules. Microbes consume the energy released for growth and reproduction [6, 13, 23]. In Nigeria, water is deemed to be polluted if the BOD level is greater than 3 mg/L. (SON). However, Fig. 9 depicts the monthly variance in BOD as determined from the groundwater. The months of January (3.20 mg/L), February (3.42 mg/L), and June (3.20 mg/L) saw the greatest BOD levels in groundwater. However, the readings were at their lowest (0.01 mg/L) in the middle of the rainy season (July to August). Therefore, variations in rainfall are the primary cause of BOD reductions in those months. While the changeover from the dry to the wet season takes place in August, the high-intensity rainfall causes groundwater dilution and a decrease in BOD [14, 43, 67]. Higher BOD levels suggest that the groundwater is contaminated with various organic substances. This contamination may be caused by anthropogenic/mining activities, seepage of sewage waste, overuse of shrimp feeds, and other factors. Wetness in seasonality has been shown to have a greater impact on BOD than dryness [9, 68]. By lowering the amounts of dissolved oxygen, high BOD poses a major threat to a variety of aquatic species [1, 4, 69, 70]. If the amount of oxygen used in water bodies was not quickly replenished, it would result in an oxygen shortage in the aquatic ecosystem. Hydrogen peroxide (H_2_O_2_) and aeration are also efficient ways to reduce excess BOD in groundwater [5, 10].

**Fig. 9:**
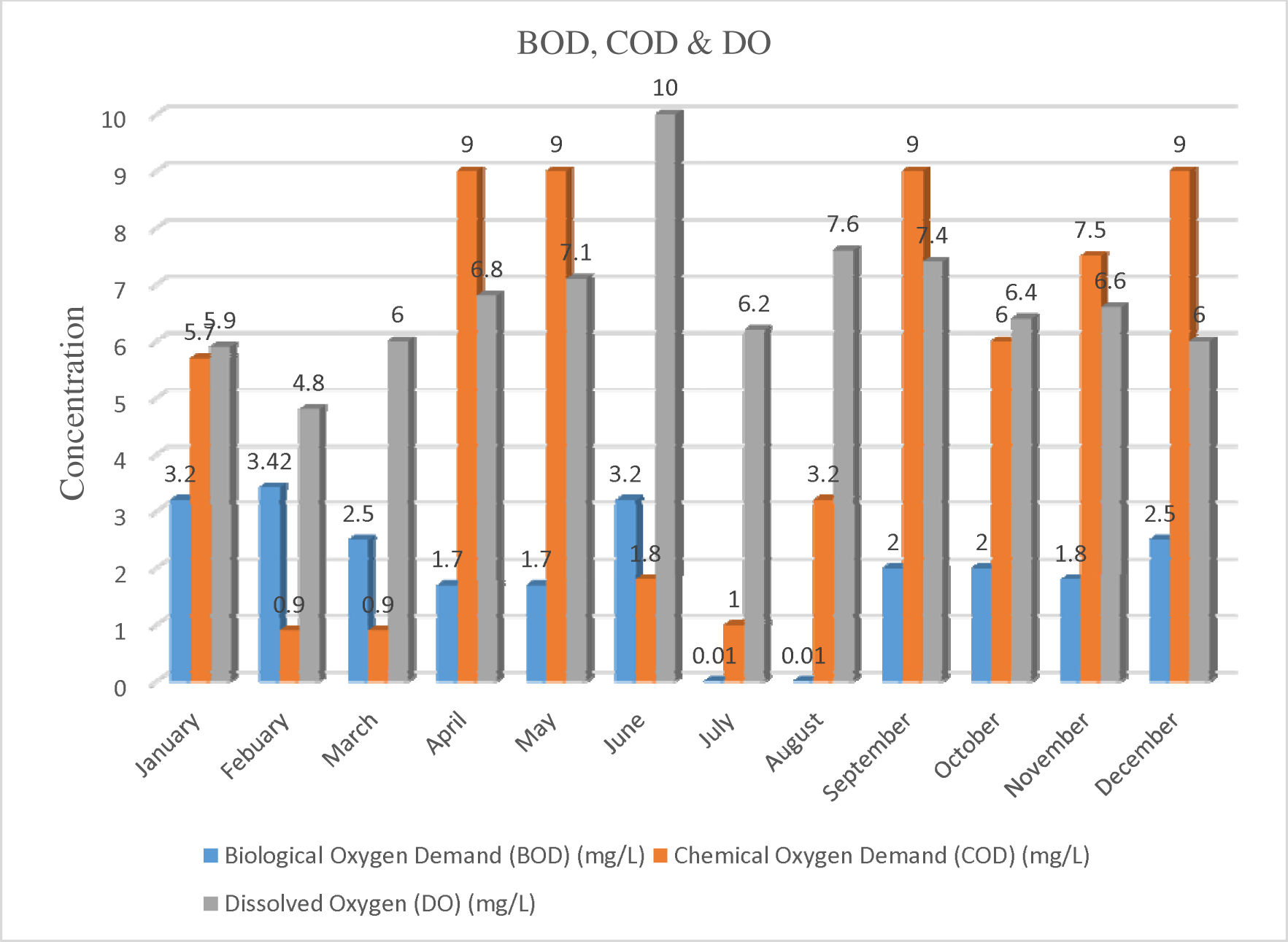
Monthly variation of BOD, COD & DO in ground water

The amount of oxygen needed for chemical oxidation in organic materials is measured by the term “chemical oxygen demand” (COD). Due to a high rate of microbial breakdown, elevated COD levels cause an oxygen deficit. It has negative effects on aquatic life. As a result, Fig. 9 presents the chemical oxygen demand in groundwater. The months of April, May, September, and December have the greatest groundwater COD levels (9.0 mg/L), while February, March, and July have the lowest values (0.9 mg/L, 1.0 mg/L, and 9.0 mg/L, respectively). As opposed to the dry season, the chemical oxygen demand (COD) is often higher during the rainy season [6, 13, 71]. The acceleration of BOD_5_, COD, phosphates, and total dissolved solids due to rainfall has a major impact on ground water pollution [7, 43, 72]. Sedimentation tanks can be used to reduce the amount of COD in groundwater while utilizing coagulants and flocculants to bind sludge together in huge masses and filter them out of the tank. Hydrogen peroxide (H_2_O_2_) can also be used to oxidize the COD and BOD levels in contaminated groundwater. A thorough feasibility analysis on the technical, economic, and environmental aspects of the under consideration groundwater, however, is very necessary before choosing the optimal approach [10, 12].

Dissolved gases, including oxygen and carbon dioxide, are important components of the freshwater system that are influenced by a number of other variables, including temperature, the partial pressure of the gases involved, respiration, salinity, and photosynthesis. While dissolved oxygen (DO) content is a term used to describe the quantity of oxygen in water. Since oxygen is essential for life, DO stands out among the other water quality indicators since it directly affects how long aquatic biodiversity will persist. In other words,

DO can be thought of as the best indicator parameter for the level of pollution in water bodies. As a result, it is necessary to use a good dynamic model to comprehend the scope of control that DO explains over biological processes [1–5]. In Fig. 9, the DO concentrations in groundwater are shown. In comparison to the dry season, when DO levels in groundwater are on average between 4.8 and 7.2 mg/L, the rainy season has higher average DO levels, ranging between 6.4 and 10.0 mg/L. The lowest DO was reported during the dry season beginning in February (4.8 mg/L), while the highest was recorded during the wet season beginning in June (10 mg/L). Consequently, the pH would rise as DO levels in water increased. Additionally, temperature, salinity, and ambient oxygen gas pressure are the primary determinants of the DO levels in groundwater bodies [5–13]. Results of water quality tests on groundwater samples’ dissolved oxygen characteristics are shown in Figure 9. In order to ensure the existence of aquatic life, the recommended DO threshold level for groundwater is 7.5 mg/L [4–12]. In several instances for groundwater samples, the value was below the advised limit. Additionally, the trial outcomes in the gold mining region of Osun State are noticeably worse than those of a DO experiment of a comparable nature carried out in an environment with oil and gas in Nigeria’s Niger Delta [1-5, 7-12]. The organic concentration and waste breakdown in groundwater are comparatively greater than in other parts of Nigeria due to the significant volume of home and industrial effluents. In general, dissolved oxygen levels were higher during rainy seasons than during dry ones. Therefore, it might be concluded that increased anthropogenic activity rates and seasonal wetness appear to have a greater impact on dissolved oxygen than their equivalents.

A groundwater sample’s total mineral concentration is gauged using TDS. Based on TDS, water can be successfully categorized. In Fig. 10, the TDS concentration in groundwater is shown. The TDS concentration in groundwater varied greatly. In June, groundwater reached its lowest point (0.65 mg/L), while in November, it reached its greatest point (577 mg/L). In comparison to the wet season, the level of TDS was obviously higher during the dry season. Therefore, elevated TDS may be a result of aquifer percolation, excessive agro-chemical use, or weathering of the rocks or soils. This demonstrates that the dry season had a stronger impact on the rainy season. The vast breakdown of salts, organic compounds, agricultural pollutants, and related chemicals first on surface waters and then seep underground where they sediment may be the cause of high TDS values in groundwater. After experiencing specific chemical changes, minerals like bicarbonates, calcium, chlorides, magnesium, potassium, sodium, and sulfates are soluble in water. They cause the water to taste bad and change colors while they’re doing these things. In general, an excessively mineralized water sample has too much TDS and causes more deformations in the quality of the groundwater [10–12]. Higher TDS causes the formation of hard water. The components of chlorides, potassium, and sodium are the most crucial elements in TDS. Although these ions are not present in greater quantities, their existence would have long-lasting impacts [7, 9]. Urban runoff, herbicides, fertilizers, and construction debris all contribute to urban groundwater contamination and a rise in TDS [1–12, 25–33]. Therefore, in order to assess the current sequences and create workable solutions, a thorough understanding of TDS based on legitimate prior study results is essential.

**Fig. 10:**
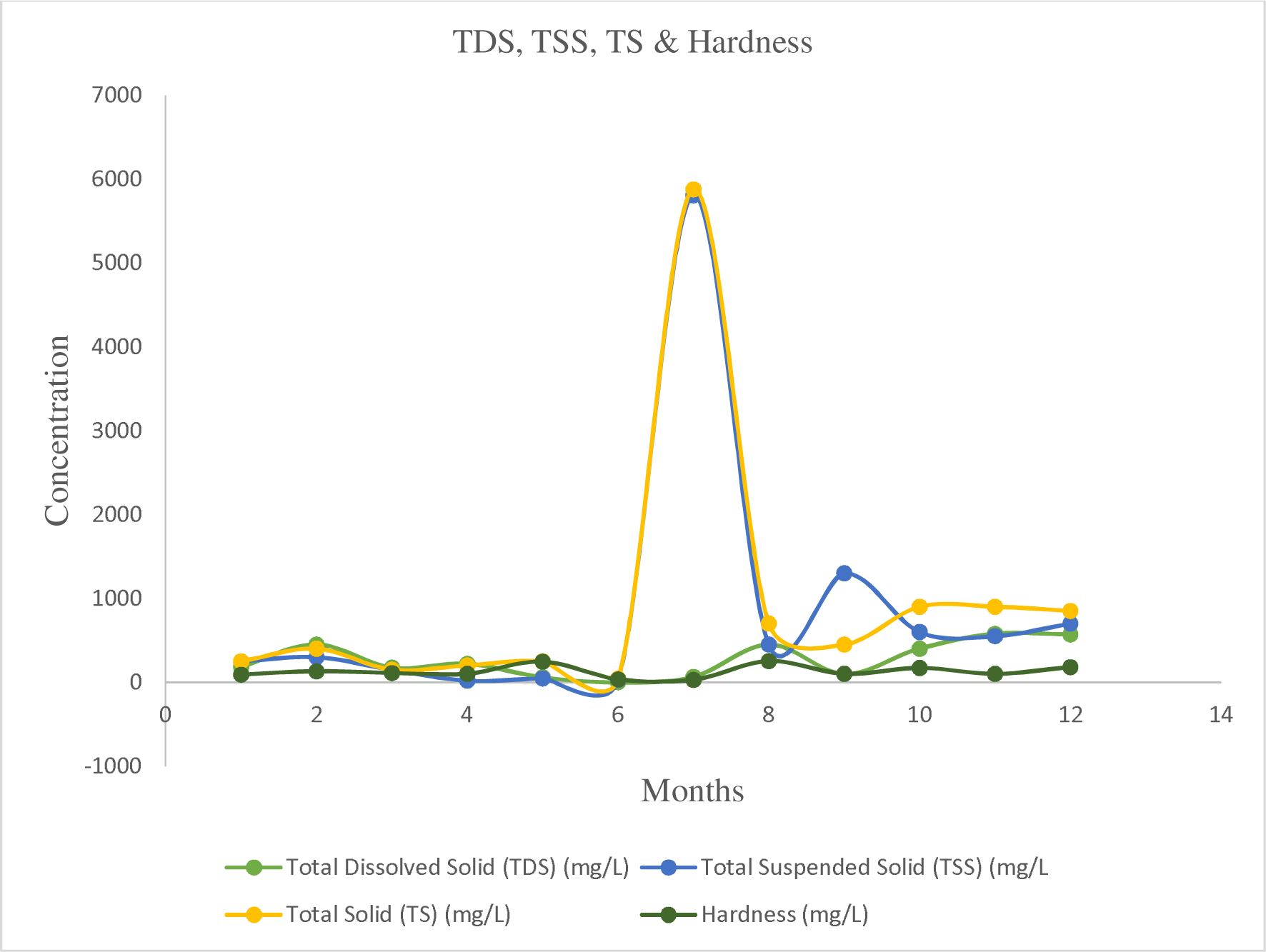
Monthly variation of TDS, TSS, TS & Hardness in ground water

The benchmark metric for measuring groundwater is TSS. It is a common contaminant that mostly encourages the growth of protozoa, sulfur bacteria, and algae cells [6, 13]. High TSS concentrations will affect water quality by raising water temperatures and lowering dissolved oxygen levels. This is because water molecules don’t absorb as much heat from solar light as suspended particles do [22]. The high rate of carbonate and bicarbonate consumption by algae as carbon sources is the cause of algal blooms brought on by higher levels of TSS. Additionally, these floating particles clog fish gills and have a negative biological impact on their immune systems and larval maturation [73, 74]. Fig. 10 displays the TSS concentrations in groundwater. TSS levels have been consistently low throughout the study’s time frame.

However, during the wet season, it peaked in the month of July at 5806.0 mg/L. The TSS ranged in value from 20.0 to 5806.0 mg/L. The lowest reading, 20.0 mg/L, was recorded in the month of April. In this investigation, all values were over the permitted upper limit. As a result, there were more dissolved materials in the research region. The extensive percolation of water through the water table during the rainy season may be the cause of higher TSS.

The levels of TS in groundwater were given in Fig. 10. The concentration of TS is generally low between mid-dry season (February) – early rainy season (June). However, the values TS inclined suddenly in the month of July (5876.00 mg/L), and levelled off in late rainy season to the early dry season. The lowest was recorded in the month of June with 42.65 mg/L.

The terrain has a significant impact on conductivity, which quantifies the concentration of charged ions in the water. Fig. 11 depicts the monthly observation of the EC level in groundwater. Particularly during the dry season and the middle to late wet season, the EC values in groundwater were greater. The highest and lowest marks were recorded in the months of November and December, respectively (800 and 780 S/cm), and 88.1 and S/cm, respectively, in May. It was noticed to be a little higher in the months of February and August (620 and 610 S/cm, respectively). As a result, the value of EC tends to rise as a result of natural groundwater recharge processes and agrochemical percolation [75]. While the WHO’s maximum allowed limit of 300 S/cm was met, all values for the Standard Organization of Nigeria (SON) under examination did not meet the restriction of 1000 S/cm. Therefore, increased conductivity readings in the months of February, August, October, November, and December could be explained by the presence of dissolved salts and other organic resources.

**Fig. 11:**
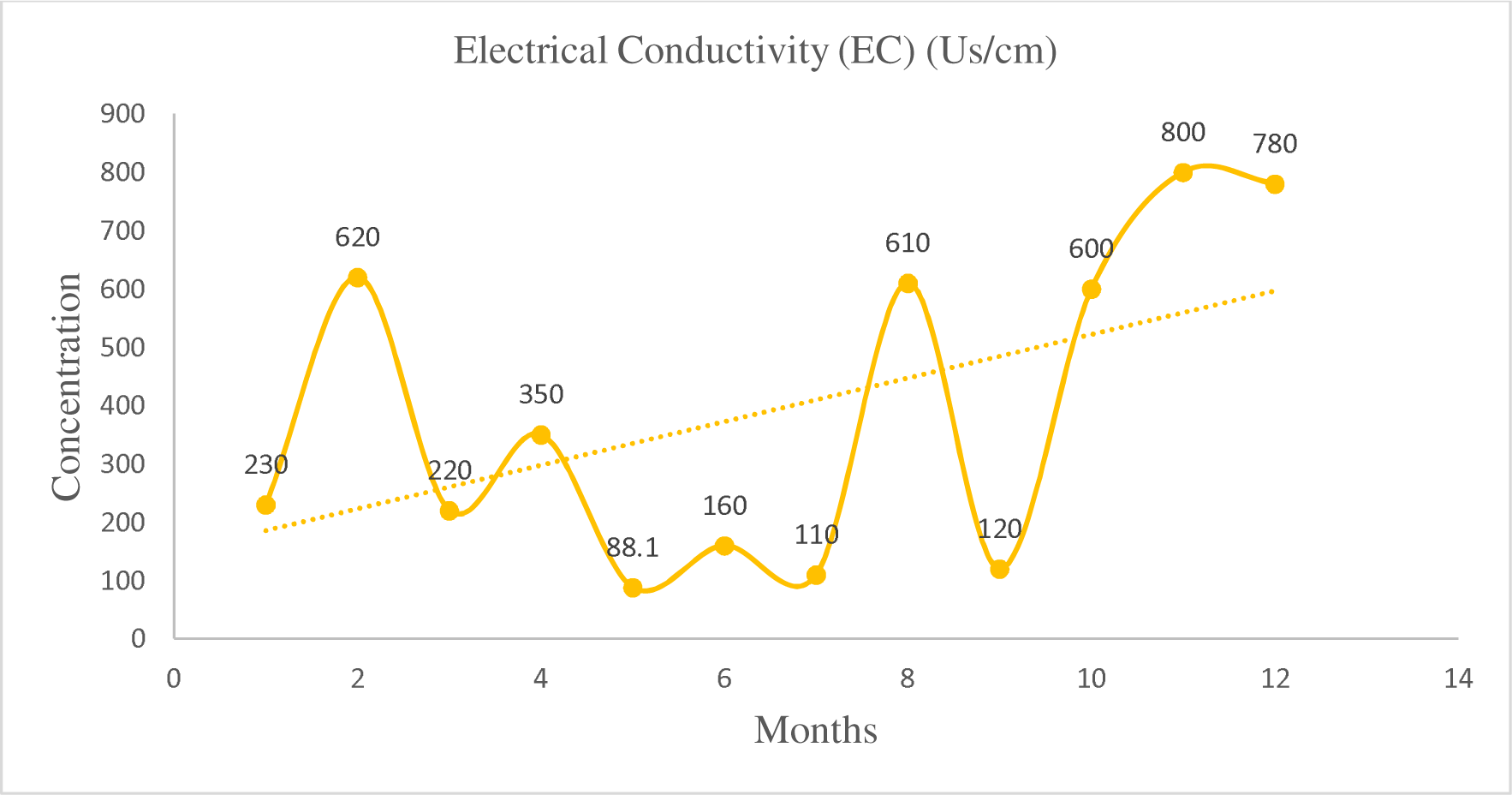
Monthly variation of Electrical Conductivity in ground water

Fig. 12 depicts groundwater turbidity. September (42.18 NTU) had the highest groundwater turbidity, followed by October (26.18 NTU), December (25.85 NTU), and January (25.85 NTU) (25.52 NTU). In May, it hit its lowest point (0.01 NTU). Groundwater turbidity ranged from 0.01 to 42.18 NTU. The varied seasonal change patterns in the respective seasons, which produce diverse sediment deposits, are to blame for the difference in groundwater turbidity values. The amount of suspended particles in groundwater determines its turbidity [76]. By measuring turbidity, it is possible to determine the extent of light penetration through water as well as the groundwater’s ability to reflect light. To determine the capacity of groundwater to release waste associated with colloids, a turbidity test is performed [77]. Turbidity is influenced by the presence of suspensions that contain clay, organic matter, planktons, silt, and other associated particles [78, 79]. These particles are released into groundwater as a result of numerous industrial and agricultural operations. As a result, they act as breeding grounds for microbes, necessitating fast disinfection in order to maintain a steady flow of groundwater for the survival of aquatic species [80]. Therefore, increasing turbidity causes suspended particles to act as adsorption sources for heavy metals including cadmium, chromium, lead, and mercury as well as agricultural pesticides and even organic substances like polychlorinated biphenyls (PCBs) and polycyclic aromatic hydrocarbons (PAHs) [81]. As a result, assessing the water turbidity shortly after a rainfall could reveal the entry of a new pollutant into a groundwater [1-8, 79]. If groundwater is used for drinking water, it must be measured for turbidity on samples every three to four hours with the utmost caution [76]. In class II and class III water, the tolerated limit for turbidity is often less than 10 NTU [9–13]. Less than 5NTU of water turbidity are visible in glasses and are intolerable owing to aesthetic reasons [23, 79]. By impeding the disinfection process by devouring the disinfectants and shielding the microorganisms, high turbidity encourages disease activity. Therefore, by treating the groundwater with chemical flocculants, turbidity must be kept below the permitted limits.

**Fig. 12:**
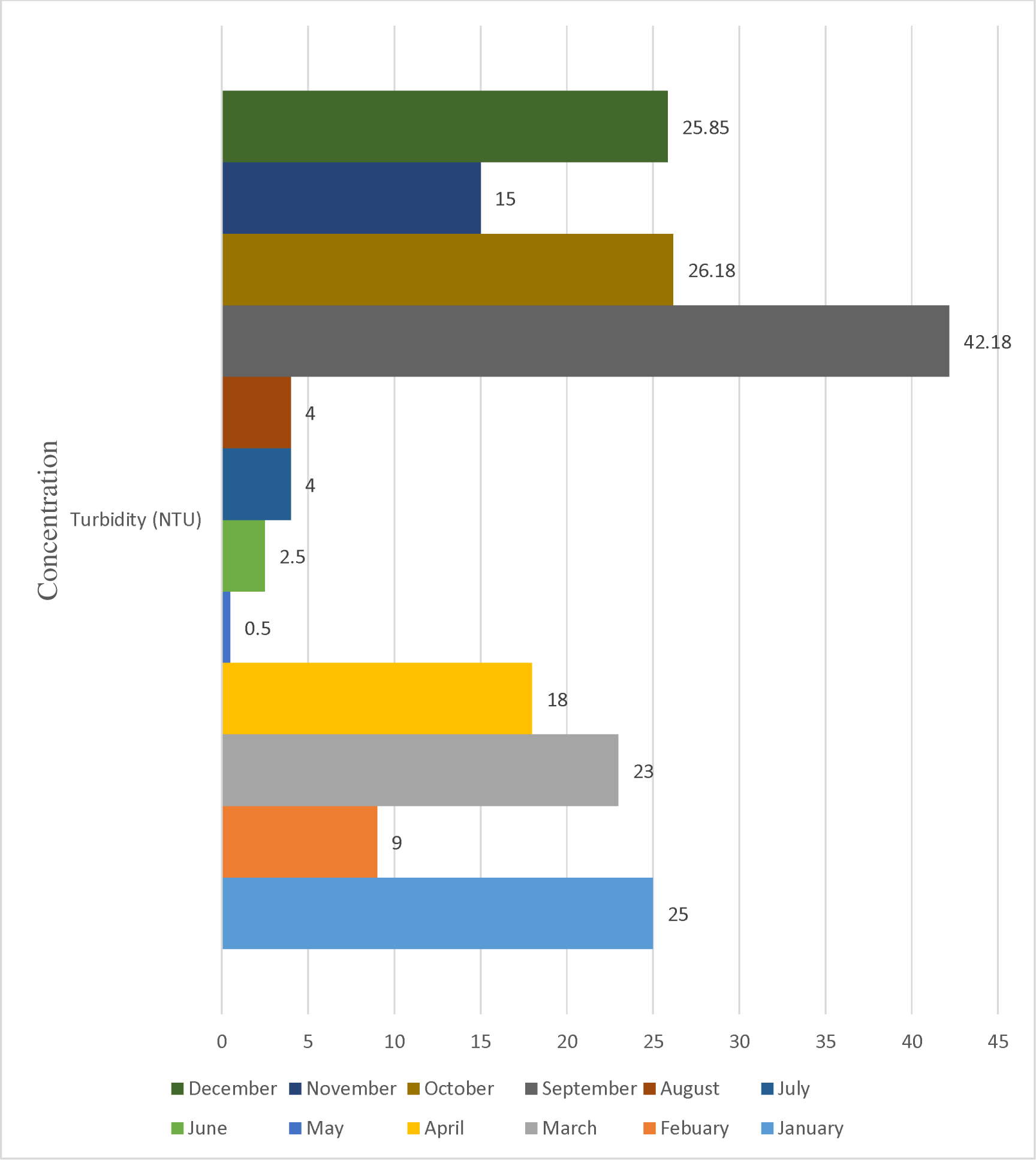
Monthly variation of Turbidity in ground water

A key factor in determining the quality of groundwater is pH. It is one of the main elements that guarantees the persistence of aquatic organisms. The concentration of active hydrogen ions in water is gauged by pH. The ideal pH range for groundwater is 7.0; anything below that range is considered alkaline, and anything above it is considered acidic [1–5, 9–12]. Both natural and man-made variables can have an impact on the pH of groundwater. The majority of natural changes are the result of interactions with nearby rocks, especially carbonate and bicarbonate forms, and other substances. Acid rain, wastewater, household garbage, and mining discharges can all cause pH to change.

Additionally, pH levels can be impacted by CO_2_ levels and the breakdown of organic materials [23, 35]. As a result, Fig. 13 illustrates the pH levels in groundwater. Throughout the dry season, pH values stayed rather constant and ranged from 3.33 to 6.8. In June and July, pH values, however, fell to a low of 3.33. In the months of August (6.05), September (6.18), and October, it grew even more (6.85). The pH levels in the samples changed according to the dry and wet seasons, showing that the samples were neutral to slightly alkaline during both. In the months of June and July, the pH values of the samples are below the permitted levels. Therefore, the pH may have increased due to the preponderance of carbonates (HCO_3_) [38]. The ideal groundwater for irrigation is typically thought to have a pH of between 7.0 and 8.0. Therefore, previous research has indicated that the majority of aquatic species and amphibians attached to water bodies would be unable to survive the pH ranges, specifically less than 3.0 or greater than 11.0 [81]. This is because they cannot withstand acidic or alkaline pH levels below 6.0 or above 10.0, respectively. Beyond this, aquatic animals need an ambient pH of 7 or higher to survive and maintain unhindered physiology, development, and metabolism [82, 83]. Beyond 8.5, pH would significantly contribute to the growth of algae that would kill fish [84] Furthermore, in extremely acidic water, heavy metals dissolve more readily. Because many heavy metals become significantly more hazardous when dissolved in water [9, 10], this is crucial information. When compared to the rainy season, it was found that the groundwater’s pH was higher during the dry seasons. In dry seasons, excessive pH ranges (pH > 6.0) were seen (January, February, March, October, November and December). However, the recorded pH magnitudes in the months of June and July were lower than those during the rainy season (May, August, and September). However, there would be two main causes for the anomalous pH range that was observed during the experimental recordings following a significant rainfall during the dry season. The first is the release of septic wastewater excretions and industrial effluents through runoffs [23].

**Fig. 13:**
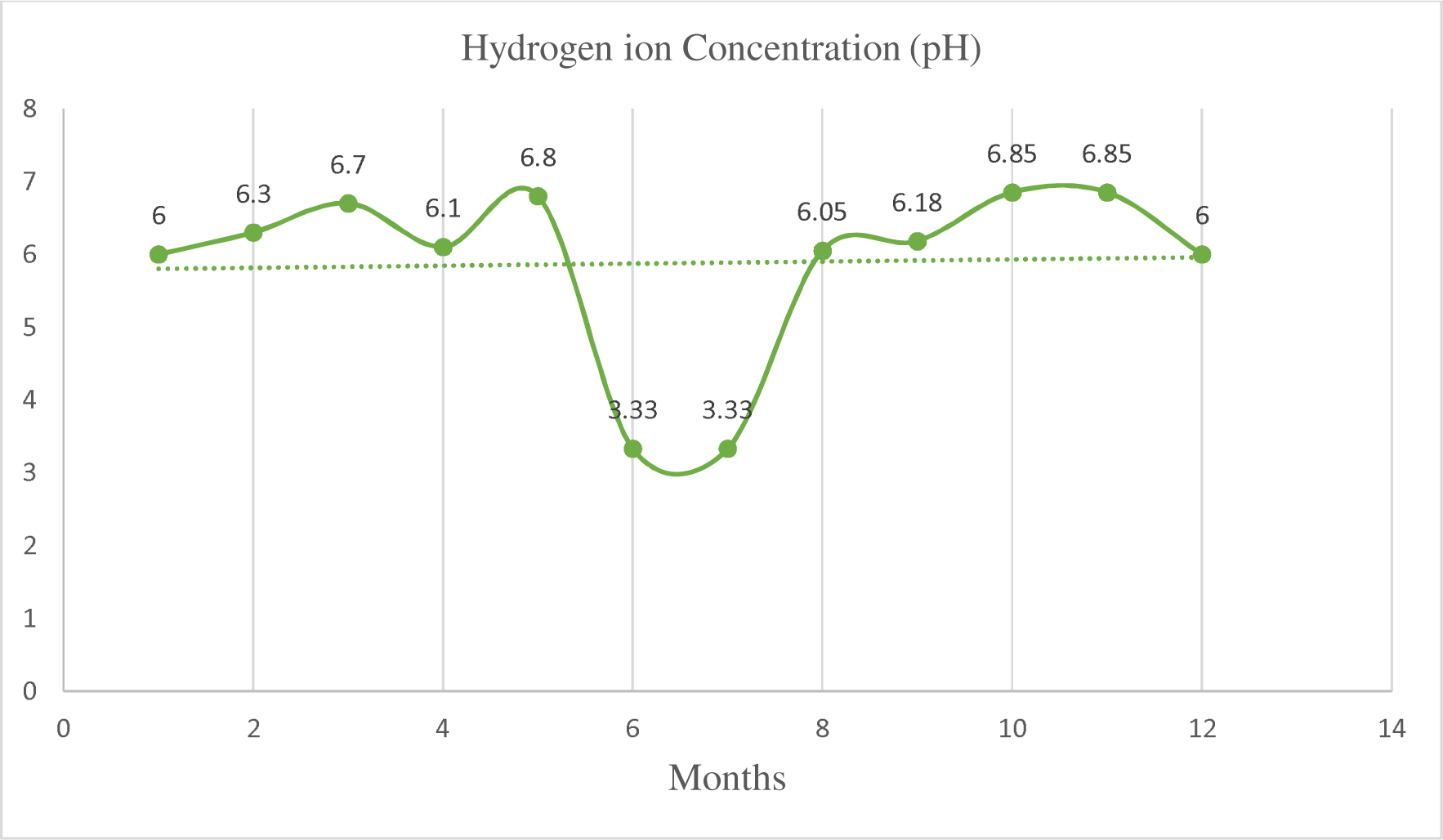
Monthly variation of Hydrogen Concentration in ground water

Hardness of ground water is frequently used to determine the degree of mineralization [12]. The main causes of ground water hardening are an increase in the concentration of calcium ions (Ca^2+^), magnesium ions (Mg^2+^), or an increase in both ions [9–12]. These metal ions have frequently caused significant hardness in both surface and groundwater during rock formation, especially during the deposition of limestone close to water sources. Bicarbonates, chlorides, nitrates, and sulfates are typical forms of these ions [76]. In addition, ions like barium (Ba^2+^), strontium (Sr^2+^), and iron (Fe^2+^) little affect the hardness of the water. As a result, Fig. 10 depicts the hardness of the groundwater in relation to the study’s months. The wet season’s peak and lowest points were found in May (245.08 mg/L) and August (252.01 mg/L), respectively, indicating that there were significant changes during this time. The lowest values were found in June (33.93 mg/L) and July (28.00 mg/L). Hardness levels above 100 mg/L are too high for drinking water. Water that is hard to drink contains a lot of HCO_3_, which makes it salty. Hard water also causes more CaCO_3_ to encrude on the soil layer, which reduces the soil’s porosity. The deterioration of humus and root respiration in the topsoil increase soil CO_2_ levels (g). Increased soil CO_2_ speeds up the breakdown of feldspar and carbonate minerals, which results in high groundwater alkalinity [9, 11]. There are currently two different categories of water hardness, including: Constant toughness. Carbonates and bicarbonates cause temporary hardness, but chlorides and sulfates cause persistent hardness [4, 12]. The total calcium and magnesium concentrations that are occupied by water are typically reported in terms of mg/L CaCO_3_ to indicate water hardness [85]. Hardness in ground water can also be brought on by the presence of hazardous elements such as arsenic, cadmium, lead, and nitrates [86]. However, because it contains a variety of dissolved salts, groundwater is thought to be quite hard.

Temperature has a significant impact on the biological activities in groundwater. Viscosity, solubility, palatability, odor, chemical reactions, and the biosorption of dissolved heavy metals are all influenced by it [1–5]. In groundwater habitats, it controls the metabolic processes, development, reproduction, distribution, and movement of aquatic creatures [9–12]. An increase in groundwater water temperature is caused by the urban heat island effect, hot water outflow from industry, and abrupt climate changes [87, 88]. As a result, Fig. 14 provides the groundwater’s temperature. Throughout the whole study period, there was no discernible change in the temperature of the groundwater. The values ranged from 24.0 to 35.0°C. During the month of June (35.0mg/L), a definite temperature peak was noted. Additionally, a spike in temperature could lead to an algae bloom, which would lower the water’s oxygen content and kill aquatic life [89]. It is claimed that warm water has lower quantities of dissolved oxygen than chilly water. Furthermore, when exposed to temperature increases, some chemicals become more harmful to aquatic life [9–12]. Thus, it appears that seasons have more of an impact on groundwater temperature than anthropogenic activity.

**Fig. 14:**
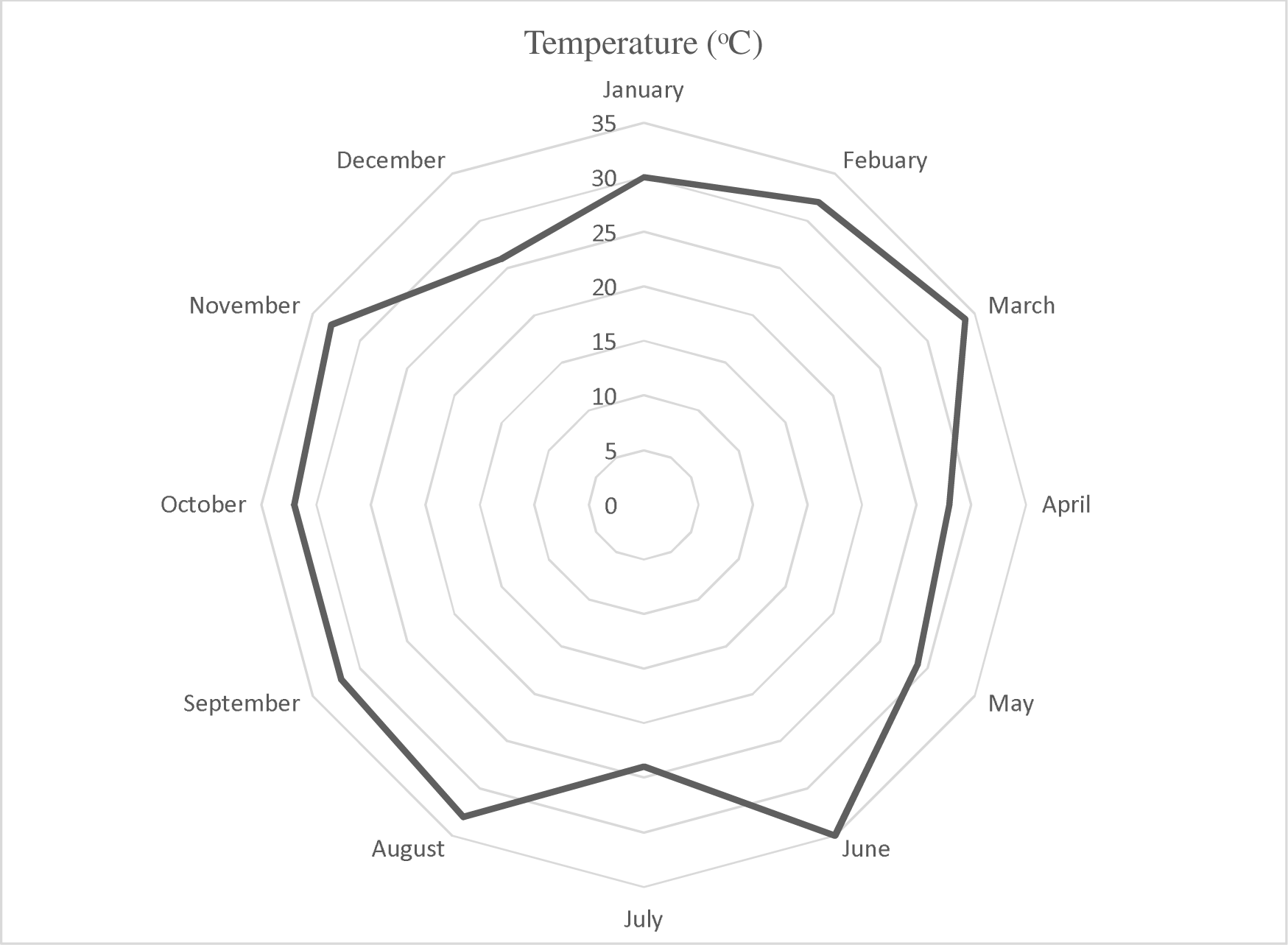
Monthly variation of Temperature in ground water

#### 3.1.1 Monthly variation in free radicals of groundwater

The calcium (Ca^2+^) monthly variation in groundwater is presented in Fig. 15. The Ca^2+^ content of groundwater was higher, particularly from mid-rainy season to mid-dry season with a peak value of 71.77 mg/L in the month of October and lowest points between June and July (0.557 – 0.926 mg/L). During the dry season it was high in the month of December (68.87 mg/L). Thus, dry season tended to influence higher concentrations of calcium than during the rainy season. The reason for the trend could be partly due to the soils and mining activities taken place in the study area as well as groundwater having deposits of calcium as natural resources and partly due to reduction in the quantity of groundwater during season to effectively reduce calcium concentrations unlike during rainy season.

**Fig. 15:**
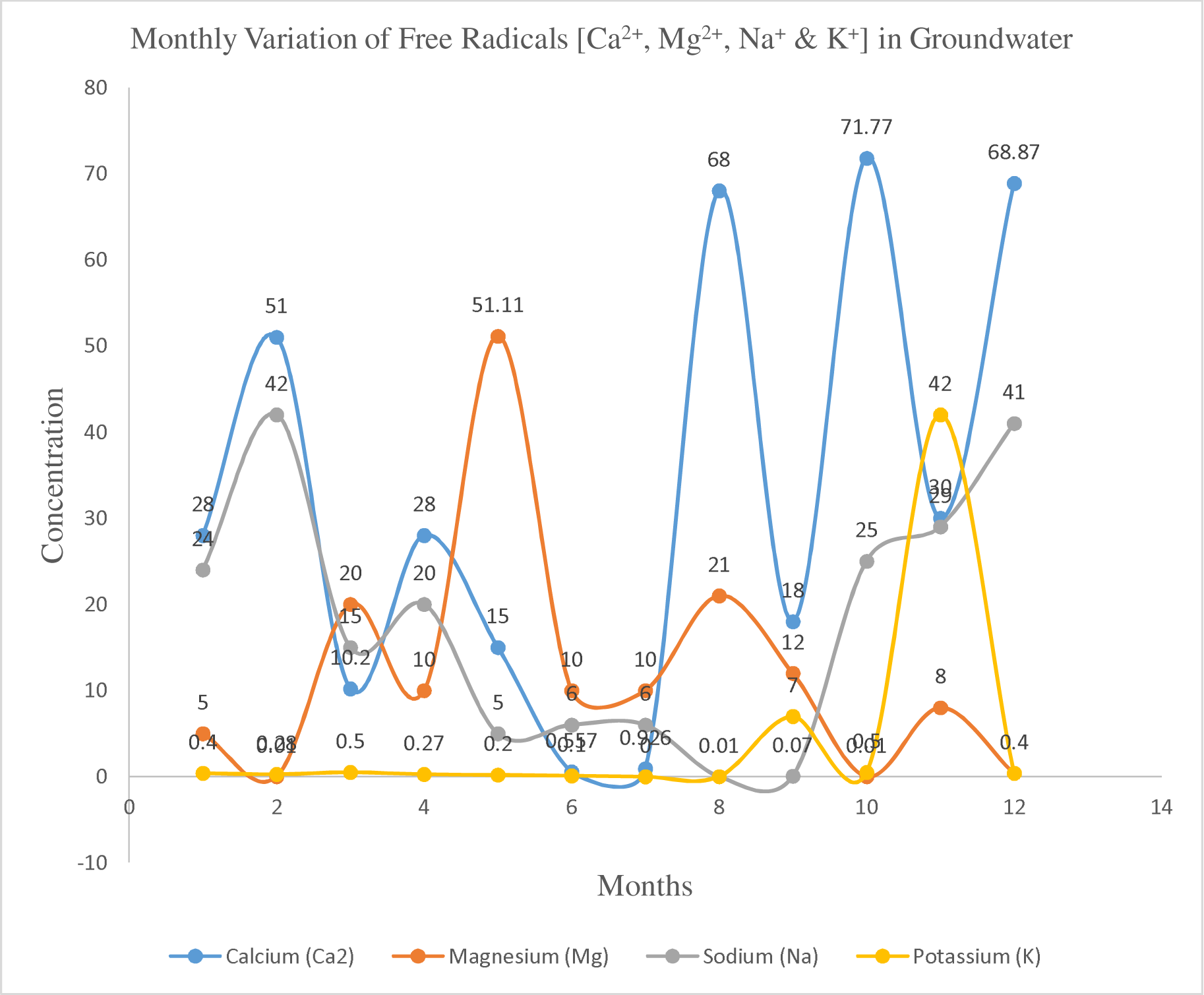
Monthly variation of Ca^2+^, Mg^2+^, Na^+^, & K^+^ in ground water

Variation of magnesium ion (Mg^2+^) concentrations in groundwater are given in Fig. 15. A clear peak was observed in groundwater at the start of the rainy season in the month of May with Mg^2+^concentration of 51.11 mg/L; while the lowest points were recorded in the dry season, precisely February (0.01 mg/L). The possible sources of Mg^2+^ in hard rock regions are mainly mica, gypsum, magnesium calcite, dolomite, and ion exchange [9-12]. The Mg^2+^ concentration may be due to the sequestration of calcium after reaching super saturation state, dolomite weathering, and excess usage of chemical fertilizers and pesticides [25-33]. Thus, it could be stated that rainy season influenced higher concentrations of magnesium than dry season. Higher values of magnesium could be attributed to roack weathering which could disintegrate chemical substances like magnesium into surface water bodies from where they percolate into groundwater.

Concentration of sodium ion (Na^+^) in groundwater are given in Fig. 15. The groundwater had higher Na^+^ in the dry season, were it reached a peak of 42.0 mg/L in February followed by the month of December with Na^+^ concentration of 41.0 mg/L. Concentration of Na^+^ was low in the rainy season compared to the dry season. The lowest point was recorded in the month of August (0.01 mg/L). Hence, higher sodium level could be an indication of natural resource deposit which later could disintegrate through chemical weathering. All values recorded in this study were below the maximum permissible limit of 200mg/L for drinking water. Thus, the Na^+^ is a salinity indicator that mainly comes in groundwater through plagioclase feldspar weathering, halite and clay minerals, and ion exchange processes. Apart from these geochemical sources, it can be present in water by domestic waste and agriculture activities.

The potassium (K^+^) in groundwater are presented in Fig. 15. The K^+^ in groundwater were generally lower than 10 mg/L in both dry and rainy seasons, except at onset of the dry season in the month of November (42.0 mg/L). The level of K^+^ in groundwater in the rainy season is lower compared to the dry season. The range values of K^+^ concentration in the groundwater is found between 0 – 42 mg/L throughout the period of the study. Higher values of potassium in the month of November could be due to the use of potassium fertilizers by farmers and which later settle underground to percolate into groundwater’s. Also, potassium could be a natural resource within the study area.

The chloride (Cl^-^) level in groundwater is given in Fig. 16. The concentration of Cl^-^ in groundwater during the dry and rainy season was low. However, Cl^-^ in groundwater increased in June (141.8 mg/L) and remained unchanged in July before plummeting in August (42.54 mg/L) and remained low for the rest of the season. All values recorded in this study were below the maximum permissible limit of 200mg/L for drinking water. The very low valu of chloride in groundwater could be attributed to increased neutralization reactions by dissolved alkaline hydroxyl containing agents. Thus, the dominancy of Cl^−^ in the month of June and July insinuates the role of evaporites dissolution and anthropogenic inputs in the hydrochemistry of groundwater. Dolomite weathering also contributes to the high alkaline earth metal concentration in a few samples. Hence, higher chloride concentrations during the rainy season may arise because of leaching from chemical fertilizers on agricultural soils, the chloride being carried into the groundwater which is the source for all reservoirs. The chloride concentrations recorded are similar to those reported in Raimi *et al.* [9], Olalekan *et al.* [10], Raimi and Sawyerr [11], Raimi *et al.* [12]. All chloride concentrations were below permissible limits (200 mg/L) for drinking water.

**Fig. 16:**
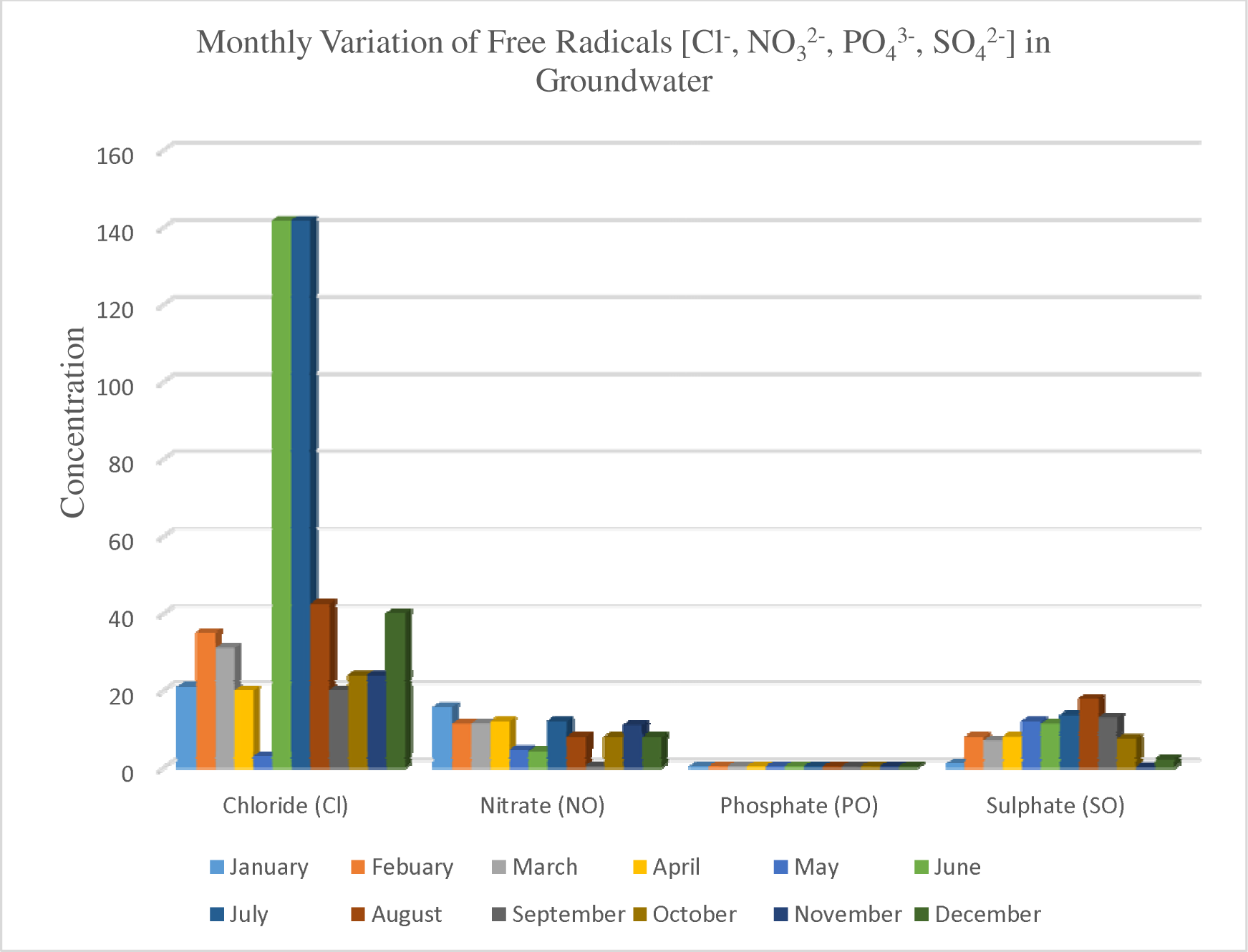
Monthly variation of Chloride, Nitrate, Phosphate & Sulphate in ground water

Due to the creation of greenhouse gases, nitrate pollution would result in increased carbon emissions and soluble chemicals that are washed into ground water. It is the main reason why algal blooms occur. Nitrates have a maximum contamination limit of 10 mg/L [10]. Fecal matter, agricultural fertilizers, industrial wastewater, solid waste dumps, and septic tanks are all ways that nitrates enter groundwater [79]. Gray-colored water that has been polluted with nitrogen affects aquatic organisms’ health and causes weariness, dizziness, and heart issues [12]. So, NO_3_^2-^ concentrations in groundwater are depicted in Fig. 16. Except for May (4.63 mg/L), June (4.20 mg/L), and September, when it dropped to its lowest (0.51 mg/L), NO_3_^2-^ concentrations in groundwater were much greater throughout the dry season and several rainy season months. It peaked in January (15.74 mg/L), when it was highest. The study’s findings were all below the 50 mg/L maximum allowable level set by national and international regulations. This pattern might also be explained by the seasonal use of nitrate fertilizers. The very low level of nitrates in the study area’s rainwater and the ensuing higher levels in ground waters may be explained by the presence of nitrogen-fixing bacteria that move atmospheric nitrogen into the soil. Therefore, home waste, agricultural effluents, and municipal waste are the main sources of nitrates. Most domestic and industrial discharges in the mining environment of Ondo state are not treated before being released into the environment [17, 22]. Increased nitrate levels hasten groundwater eutrophication. However, the transformation of nitrates into nitrites results in a number of health effects for people, including thyroid, cancer, and acute illnesses, as well as blue baby syndrome (also known as methaemoglobinaemia) for newborn newborns [7–11]. Current newspaper articles on community opinions, study findings on the central Niger Delta waterways provide circumstantial support for categorizing recent groundwater nitrate pollution problems as a serious danger [90]. There are a number of methods that can be used in the mining environment of Ondo state to reduce nitrate pollution, including the use of bioreactors, drainage water recycling, and controlled drainage. It is crucial to choose and implement the most appropriate method to reduce current nitrate pollution trends in the state. In conclusion, the variations in nitrate concentration between the seasons were visible in the reservoirs. Nitrite levels in groundwater were greater during the dry season. The measured nitrate amounts, however, were far below the 50 mg/L drinking water limit that is advised [65, 66]. It should be highlighted that these results are significantly lower than the range of 0.3 to 7.0 mg/L obtained in studies carried out in an oil and gas environment [1-5, 8-12].

Pollution sources of phosphates in groundwater include agricultural runoff, detergents, and domestic and industrial wastewater effluents [79]. The maximum allowable phosphate concentration for ample aquatic life is in the range of 0.1 - 0.3 mg/L [13]. Higher phosphate content would cause eutrophication, which gives both short-term and long term BOD in groundwater. The maximum phosphate limit that a groundwater could buffer without eutrophication is 0.1 mg/L [10, 12]. Phosphates should not be completely negligible in ground water scenarios since they have an immense contribution to the growth of root systems of aquatic plants. On the other hand, phosphates contribute more than nitrates to algal blooms [76]. Thus, PO_4_^-^ concentrations are given in Fig. 16. A similar trend of PO_4_^-^ levels was observed in groundwater in the dry and rainy season for most part of the study period. However, the highest concentration of PO_4_ ^-^ was observed in the month of February and November with the average mean of 0.33 mg/L. Furthermore, it is also observed that the proper treatment of groundwater, phosphorous removal is generally facilitated using the chemical precipitation method by incorporating tertiary filtration and metal salts.

Sulphate (SO_4_^2-^) in ground water can be measured using turbidimetric tests and are prominent contaminants in general water bodies. The leaching of natural deposits, atmospheric deposition, and anthropogenic discharges directly influences the concentration of sulphates and their complex consolidation [9-12]. In general, sulphates are discharged into bodies of water by industries such as mines, textile mills, smelting operations, Kraft pulp, tanneries, paper mills, agricultural run-off, and sewage [91, 92]. However, high sulphate concentrations are of particular concern to the mining industry [93]. Thus, SO_4_^2-^ level in groundwater during period of the study is presented in Fig. 16. Groundwater SO_4_^2-^ levels was low at the start of the dry season (November – January) (0.21 – 1.01 mg/L) but increased steadily from the mid-dry season (February) into the raining season reaching a peak of (17.92 mg/L) in August. It started declining in the late rainy season, September (12.99 mg/L), reaching its lowest point in November (0.21 mg/L). While, in nature, sulphates exist as barites (BaSO_4_), gypsum (CaSO_4_.2H_2_O) and epsomites (MgSO_4_.7H_2_O) and the interaction of water in those forms would contaminate sulphates with water bodies [12, 94]. Compounds such as Na_2_SO_4_, K_2_SO_4_ and MgSO_4_ are extremely soluble in water, whereas CaSO_4_, BaSO_4_ and many heavy metal sulphates are less soluble in water [12]. An increasing amount of sulphate can cause chronic effects on aquatic organisms [13, 91]. The sulphate concentration of ground water should be lower than 100 mg/L because seawater contains about 2,700 mg/L of sulphate [91]. Summarily, there were large differences in sulfate concentration between seasons. The main difference was found when dry season water was compared with rainy season. The main source of sulfate in the water is mineral dissolution. Sulfate in water causes noticeable taste changes. The taste threshold ranges from 250 mg/L for sodium sulfate to 1,000 mg/L for calcium sulfate [12]. High sulfate concentration can cause permanent hardness, but levels were below the WHO’s maximum acceptable level for drinking water (250 mg/L) in groundwater reservoirs [9-12]. A study in the Niger Delta also reported sulfate concentrations in the range 11 to 26 mg/L in line with this study [1-5, 7-12].

#### 3.1.2 Monthly variations in heavy metal concentration in groundwater

High toxicity, persistence, and bioaccumulation are traits of heavy metals [7– 13]. They can persist in soils for a very long time and build up in the food chain, which poses a serious threat to the environment of the soil, the security of the food supply, and human health [17]. While the primary causes of the heavy metal poisoning of groundwater are human activities. Due to the harmful chemicals utilized in all those circumstances, agricultural and industrial effluents could hasten the contamination of ground water with heavy metals [23, 35]. Due to their biological nature, heavy metals accumulate in groundwater in high concentrations. Due to anthropogenic pressure, environmental toxicity of heavy metals has resulted in poor water quality and the loss of aquatic habitats, which has resulted in the depletion of resources worldwide [47–49]. Heavy metals are also non-biodegradable and carcinogenic, thus their presence in large levels in water could have detrimental effects on the health of living things [6, 13, 47–49]. Fish that are exposed to heavy metals develop abnormally, die, and have delayed hatching [6, 13]. Arsenic, cadmium, lead, mercury, zinc, nickel, copper, and chromium are the heavy metals that are most frequently found in groundwater [7–12, 19, 20]. The kind, compound formation, and amount of deposition of heavy metals can all affect how harmful they are [95]. Groundwater contamination by heavy metals is currently a significant environmental problem. Heavy metals build up in various plant tissues and interfere with many metabolic processes, such as photosynthesis and respiration inhibition and degeneration of major cell organelles, which lead to stunted growth, delayed germination, chlorosis, premature leaf fall, senescence, decreased crop yield, and loss of enzyme activities [96, 97]. As a result, heavy metals affect the growth and development of plants. Ingestion of heavy metals like Cd and Zn in human’s results in numerous illnesses such acute gastrointestinal, musculoskeletal, and respiratory problems as well as harm to the brain, heart, and kidney.

In many places of the world, the widespread use of lead has resulted in significant environmental damage and health issues. Lead is a highly poisonous metal. The primary industrial operations, food and smoking, drinking water and residential sources are the sources of lead exposure. Fuel and house paint were the main sources of lead, but it has since spread to plumbing pipes, pewter pitchers, storage batteries, toys, and faucets [98]. While sources of lead contamination include mining, paint, battery waste, coal burning, pesticides, herbicides, and emissions from the burning of leaded gasoline. Fig. 17 shows the changes in Pb concentration in groundwater during the course of the study’s month. Pb levels in groundwater reach their highest point in December (2.11 mg/L). However, groundwater reached its lowest point in July (0.32). The Pb concentration measurements for the months of February (1.30 mg/L), March (1.07 mg/L), April (1.04 mg/L), and May (1.30 mg/L) deviate somewhat from one another. In contrast to other metals like zinc, copper, and manganese, lead is an exceedingly poisonous heavy metal that interferes with a number of physiological processes in plants but has no biological purpose. A plant exposed to high levels of lead speeds up the creation of reactive oxygen species (ROS), which damages lipid membranes and, in turn, damages chlorophyll and photosynthetic activities, slowing down the growth of the plant as a whole [99]. According to some research, lead can retard the growth of tea plants by lowering biomass and degrade the quality of tea by altering the components’ qualities [100]. Lead treatment has been shown to significantly alter plant ion uptake, even at low concentrations, which in turn causes large metabolic alterations in photosynthetic capability and eventually results in a considerable suppression of plant growth [101].

**Fig. 17:**
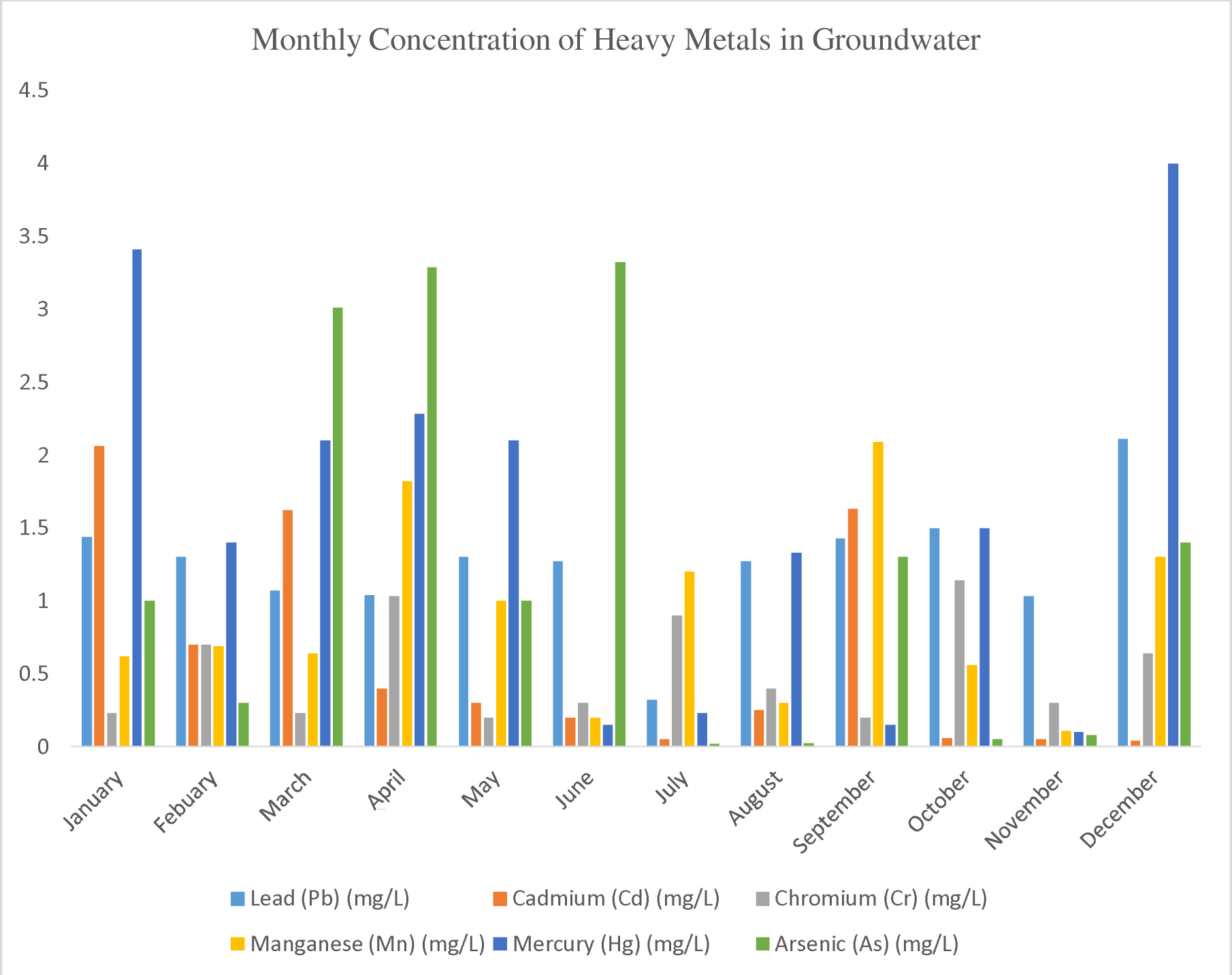
Monthly variation of Pb, Cd, Cr, Mn, Hg & As in ground water

Humans can get acutely and chronically intoxicated after being exposed to Cd metal, mostly by ingestion and inhalation. For many years, cadmium that has been dispersed in the environment will persist in soils and sediments. The concentration of cadmium (Cd) in groundwater is shown in Fig. 17. In comparison to the wet season, the midpoint of the dry season saw greater Cd levels in groundwater. The highest groundwater Cd values were recorded in January (2.06 mg/L), and the lowest readings were recorded in July (0.051 mg/L). The month of September (1.63 mg/L) saw the only elevated Cd concentration during the wet season. The harmful effects of Cd include Itai-Itai disease, carcinogenicity, mutagenicity, renal and lung damage, weak bones, and gastrointestinal disorders [10, 12].

Chromium is anthropogenically introduced into the environment through fertilizers and sewage [13, 102]. Chromium (Cr) concentrations in groundwater are depicted in Fig. 17. When compared to the dry season, the levels of Cr were greater in the rainy season, particularly in the months of April (1.03 mg/L) and October (1.14), which indicate the beginning and end of the rainy season, respectively. The result ranged from 0.21 to 1.14 mg/L. Thus, chromium is produced naturally by the burning of coal, oil, ferro cromate refractory material, pigment oxidants, catalyst, chromium steel, fertilizers, drilling for oil wells, and tanneries used for metal plating. Whilst chromium is widely used in a variety of industries, including metallurgy, electroplating, the manufacture of paints and pigments, tanning, the preservation of wood, the creation of chemicals, and the manufacture of pulp and paper. These businesses contribute significantly to the chromium contamination that negatively affects ecological and biological species [102].

Concerns regarding chromium pollution are raised by a variety of industrial and agricultural methods that raise the hazardous level in the environment. When agriculture is practiced continuously near a mining area, Cr is continuously released into the environment through Cr residues, Cr dust, and Cr waste water irrigation, causing soil pollution that disrupts the soil-vegetable system and also affects the yield and quality of vegetables for human consumption [103]. Chromium excess above the allowable limit is harmful to plants because it adversely affects their biological processes and enters the food chain when these plant components are consumed.

The concentration of Mn in groundwater are presented in Fig. 17. No clear trend was observed in Mn concentration in the groundwater. However, the concentration of Mn was at its peak in the month of April (1.821 mg/L) and September (2.09 mg/L) and reached its lowest point in November (0.11 mg/L). There is slightly difference in the values of Mn concentration in the month of January (0.62 mg/L), February (0.69 mg/L) and March (0.64). A study in the Niger Delta also reported manganese concentrations in the range 11 to 26 mg/L in line with this study [1-5, 7-12.

The naturally occurring metal mercury is a bright, silver-white, odorless liquid that, when heated, transforms into an odorless, colorless gas. Mercury is a well-known dangerous metal that is extremely bioaccumulative and toxic, and its toxicity is frequently the cause of acute heavy metal poisoning. Mercury (Hg) levels in groundwater are depicted in Fig. 17 as a result of the harmful effects its presence has on the marine environment. With Hg concentrations of 4,00 mg/L and 3,41 mg/L, respectively, December and January had greater levels of Hg, indicating that the mid-dry season has higher concentrations of Hg. During the rainy season, only the months of April (2.28 mg/L) and March (2.10 mg/L) had greater Hg concentrations. The range of values found in the groundwater was between 0.04 and 4.001 mg/L. Therefore, anthropogenic activities such farming, municipal wastewater discharges, mining, waste incineration, and industrial wastewater discharges are major sources of mercury pollution [7, 17, 104]. The three main forms of mercury are metallic elements, inorganic salts, and organic molecules. Each form of mercury has a unique toxicity and bioavailability. These types of mercury are commonly found in lakes, rivers, and seas where they are ingested by microorganisms and converted to methyl mercury before finally undergoing biomagnification, which causes severe disruption to aquatic life. The main way that humans are exposed to methyl mercury is through the consumption of these polluted aquatic animals [7–12, 17–20, 105]. Thermometers, barometers, pyrometers, hydrometers, mercury arc lamps, fluorescent lamps, and catalysts all make substantial use of mercury. Additionally, it is utilized in the pulp and paper industry, in batteries, and in dental products like amalgams.

In Fig. 17, the levels of arsenic (As) in groundwater are shown. As concentrations in groundwater ranged from 0.05 to 3.32 mg/L. When compared to the other study months, the concentration of arsenic was obviously higher in the months of March (3.01 mg/L), April (3.29 mg/L), and June (3.32 mg/L). With an As concentration of 0.05 mg/L, October’s concentration was the lowest ever. As a result, both natural geologic processes and human activity have contributed to arsenic contamination. Arsenic is produced by humans through processes like ore processing and mining.

Arsenic can be released during the smelting process into the air and soil [17, 48, 106]. Through runoff and groundwater ejection, these types of sources can have an impact on the quality of surface water. Geologic sources, such as minerals containing arsenic, are another method for ground water to get contaminated in a mining setting. Sedimentary and meta-sedimentary bed rocks constitute the third category of sources [8–12]. Arsenic is present in the majority of paints, dyes, soaps, metals, semi-conductors, and medications. Arsenic is also released in greater quantities to the environment through the use of some pesticides, fertilizers, and animal feeding practices. It has been determined that the inorganic forms of arsenic, such as arsenite and arsenate, are more hazardous to human health. They are extremely carcinogenic and can result in skin, bladder, liver, and lung cancer. Arsenic exposure in humans occurs through the air, food, and water [7, 107-110]. One of the main contributors to arsenic toxicity in more than 30 nations throughout the world is drinking water [111]. Human health may be at risk if the quantity of arsenic in groundwater is 10-100 times higher than what is recommended by the WHO for drinking water (0.01 mg/L) [7–12]. Natural mineral deposits or poorly disposed of arsenical chemicals or pesticides may contaminate groundwater. Arsenic toxicity can be acute or chronic, and arsenicosis is the medical word for persistent arsenic toxicity.

## 4. Conclusion

Industrial development is contributing to an increase in global environmental pollution. In order to comprehend the hydro-geochemistry of the groundwater and its appropriateness for drinking and irrigation in the gold mining regions of Osun State, South-West Nigeria, this study is being conducted. In order to draw the appropriate results, the current study takes into account a seasonal variance. There are significant differences in such values over the course of a year depending on whether it was dry or rainy. As a result, the corresponding alterations were explained from a logical standpoint using the conclusions from pertinent research studies. As a result, human involvement in ground water has occurred frequently, significantly degrading its quality over time. Major anthropogenic activities, such as mining, agriculture, obstruction of water for irrigation and drinking, washing of clothing and utensils, discharge of sewage water and industrial effluent, disposal of municipal solid waste along the bank of the river, and religious ritual activities along the stretch, have been posing a serious threat to biota. Effective laws, regulations, and the identification of locations with higher physico-chemical and heavy metal concentrations are thus required. Because of the negative consequences that heavy metals and physico-chemical impacts impose, failure to restrict the exposure will cause serious issues in the future. A significant step towards prevention can be taken by tracking exposure to heavy metals and possibly taking action to reduce future exposure in both persons and the environment. For the purpose of developing effective strategies to avoid hydro-geochemistry toxicity, both national and international cooperation is essential. Additionally, because this investigation has highlighted the characteristics, contributing to contamination, it will aid in future ground water control management programs. Therefore, it is essential to create a thorough ground water quality monitoring program for South-West Nigeria. By presenting a thorough roadmap of the crucial factors that must be evaluated on groundwater in a mining context, this work will significantly advance the field of science worldwide. The following are the findings of the current study:

i. The research region’s groundwater is adequate for drinking in terms of BOD, COD, Na, K, Ca^2+^, Cl, and SO_4_^2^, but it is unfit due to greater concentrations of lead, mercury, cadmium, manganese, and arsenic, as well as higher levels of color, pH, DO, EC, TDS, TSS, and TS. As a result, the analytical findings show that the study region’s groundwater is unfit for drinking. Nearly half of the samples that were collected throughout both seasons were found to be contaminated to an extreme, showing the extent of the problem in the area under study.

## Data Availability

All data produced in the present work are contained in the manuscript

## Acknowledgement

The authors are thankful to the Department of Environmental Health Sciences, Kwara State University for the encouragement of research activity and for providing necessary laboratory facilities.

## Authors’ contributions

Conceptualization, ASO and SOA.; methodology, ASO and MOR.; software, OAO.; validation, SOA, SOH and OAO.; formal analysis, MOR and OAO.; investigation, ASO and SOA.; resources, ASO and SOH.; data curation, MOR and OAO.; writing - original draft preparation, ASO, SOA and SOH.; writing - review and editing, ASO and SOA.; visualization, MOR and OAO.; supervision, SOA and SOH; project administration, SOA and SOH. All authors have read and agreed to the published version of the manuscript.

## Grant Support Details

The present research did not receive any financial support.

## Conflict of Interest

The authors declare that there is not any conflict of interests regarding the publication of this manuscript. In addition, the ethical issues, including plagiarism, informed consent, misconduct, data fabrication and/ or falsification, double publication and/or submission, and redundancy has been completely observed by the authors.

## Life Science Reporting

No life science threat was practiced in this research.

## Notes

### Competing Interest Statement

The authors have declared no competing interest.

### Funding Statement

This study did not receive any funding

